# A cross-domain test battery for comprehensive hearing loss characterisation using functional, physiological, and vestibular measures

**DOI:** 10.64898/2025.12.30.25343205

**Authors:** Mareike Buhl, Shiran Koifman, Abile Serge Magbonde, Koray Koçoğlu, Sabine Hochmuth, Elie Partouche, Arnaud Coez, Andreas Radeloff, Hung Thai-Van, Sylvette Wiener-Vacher, Grégory Gerenton, Anna Warzybok, Paul Avan, Birger Kollmeier

## Abstract

**Objective:** This binational cross-centre study analyses a consented audiological-vestibular test battery for characterising age-related hearing loss, enabling precise phenotyping of suprathreshold functional, physiological, and vestibular factors beyond audibility.

**Design:** Statistical analysis of centre effects to assess comparability of the test battery measured at two centres (Germany and France); statistical analysis of age and pure-tone average (PTA) effects per test to identify potential covariates.

**Sample:** n = 55 (39 German and 16 French) participants with hearing thresholds better than the age-dependent median of the PTA, aged 40 years or older.

**Results:** Age- and PTA-dependent reference data were derived. Due to negligible centre effects, all data were pooled across centres. Age and PTA effects were identified for some tests, especially for audiological-functional tests. No age effects were found for vestibular tests.

**Conclusions:** Normative values for a clinically feasible, multidimensional audiological-vestibular test battery were provided, including several measures whose age and PTA dependencies were previously unclear. Age and PTA should be considered as covariates for interpretation of these tests in future applications such as, e.g., phenotype-genotype relations in specified cohorts.

## Introduction

Individualised characterisation of hearing function is a central aspect in audiology, as hearing loss manifests with considerable variability across patients arising from differences in severity, underlying aetiology, and other contributing factors. Comprehensive assessment not only guides personalised clinical management but also provides insights into the mechanisms underlying auditory dysfunction; however, it is limited due to time constraints in clinical settings. The current paper attempts to establish a clinically feasible test battery which has been consented in a binational cross-centre project. By providing age- and hearing-loss-related normative values for a yet limited population of participants with hearing thresholds better than the age-dependent median of the pure-tone average (PTA), the practical feasibility of this test battery is demonstrated.

Pure-tone audiometry, a gold standard in audiology, captures functional hearing thresholds, but a more complete picture requires the integration of physiological, vestibular, and even genetic information. Traditionally, additional dimensions of hearing loss have been assessed either from a functional or a physiological perspective, yet the links and complementary contributions of both domains remain unclear.

From the *functional perspective*, suprathreshold deficits such as impaired speech recognition in noise, altered loudness perception, or neural processing deficits have been proposed as key elements of a more comprehensive characterisation of hearing impairment (e.g., Kollmeier & Kiessling, 2018), and can be quantified using a range of psychoacoustic tests. These deficits have been assessed based on measurements such as speech recognition in noise, tone-in-noise detection or adaptive categorical loudness scaling (Hülsmeier et al., 2022; Hülsmeier & Kollmeier, 2022; Schädler et al., 2020; Sanchez-Lopez et al., 2018; Saak et al., 2022). Importantly, suprathreshold deficits are not always predictable from the audiogram. Recent work has further demonstrated that age effects play a crucial role in speech in noise performance, even beyond audiometric threshold elevation, as shown in O’Brien et al. (2024). Early on, Plomp (1978) proposed a useful distinction between an attenuation component, reflecting audibility loss as measured by pure-tone audiometry, and a distortion component, referring to deficits that emerge at suprathreshold levels, such as reduced speech recognition in noise or degraded sound quality. This framework underscores that hearing impairment is not a one-dimensional phenomenon but instead involves multiple perceptual domains that can be differentially affected by aging.

From the *physiological perspective*, electrophysiological tests provide insights into the underlying pathology of these functional deficits and linking them to functional tests could provide the potential for more causal diagnostics of hearing impairment (Plack et al., 2014). Similarly, a relationship between hearing loss and vestibular deficits is suspected, given the close anatomical and physiological proximity of cochlear and vestibular hair cells within the same endolymphatic compartment (Petit & Richardson, 2009). Damage to shared structures such as hair cells, the stria vascularis, or supporting cells can therefore compromise both auditory and vestibular function. Epidemiological studies suggest that vestibular hypofunction is more prevalent in individuals with sensorineural hearing loss than in age-matched controls, although the mechanisms underlying this association remain incompletely understood in some cases (Omer et al., 2024). Including vestibular testing in a comprehensive test battery thus offers the opportunity to uncover potential co-morbidities and to examine whether specific patterns of cochlear dysfunction are systematically linked to vestibular deficits. For example, vestibular impairment is reported as associated to cochleovestibular loss (Byun et al., 2020; Castellucci et al., 2023), highlighting the relevance of targeted vestibular assessments in age-related hearing loss.

Importantly, many of these functional and physiological changes are exacerbated by aging. Presbycusis, or age-related hearing loss, represents one of the most prevalent sensory deficits in the elderly population and is characterised not only by elevated thresholds in the high frequencies but also by suprathreshold deficits such as impaired temporal and spectral resolution, reduced speech recognition in noise, and cognitive-auditory interactions (Gates & Mills, 2005; Füllgrabe, 2015). Normative reference data for the expected age-related threshold elevation across frequencies have been codified in ISO 7029 (2017), providing a widely used standard for quantifying age effects on audiometric thresholds. On the physiological level, age-related degeneration of cochlear structures, neural synaptopathy, and potential comorbid vestibular decline all contribute to the multifactorial nature of presbycusis. A test battery that integrates functional and physiological dimensions of hearing may provide valuable means to disentangle the complex interplay between auditory and vestibular aging.

Several test batteries have been developed to move beyond pure-tone audiometry towards a more multidimensional characterisation of hearing function. Examples include the HEARCOM test battery (van Esch et al., 2013), the BEAR test battery (Sanchez-Lopez et al., 2021), and more recently, the Oldenburg Hearing Health Record (OHHR; Jafri et al., 2025). HEARCOM and BEAR primarily focus on functional measures, including psychoacoustic tasks, speech-in-noise performance, and spectro-temporal modulation (BEAR), but do not systematically incorporate physiological assessments. The OHHR collects large-scale behavioural and clinical data, including audiometry and self-reported outcomes, but currently lacks integrated physiological measurements. Qian et al. (2021) used perceptual and physiological tests to assess aging effects in normal-hearing listeners, enabling a more comprehensive cross-domain evaluation. In summary, these test batteries illustrate the spectrum of approaches for characterising hearing function, from functional-only to combined functional-physiological assessment, highlighting the need for integrative frameworks that capture the full complexity of auditory and vestibular deficits.

Taken together, several domains may be important for comprehensive hearing loss characterisation beyond audibility, but relationships between domains and tests remain unclear due to insufficient data availability (or the unmet need of a test battery comprising all domains). To address these gaps, the PRESAGE (Towards PREciSion audiology for AGE-related hearing loss) project aims at providing such a comprehensive characterisation of hearing loss across multiple domains. The selection of tests to be performed in clinical settings (with the typical clinical time restrictions necessitating a time-comprehensiveness compromise) is based on a multidisciplinary consensus across the project team. The project involves data collection at the University of Oldenburg (UOL; Germany) and at the Institut de l’Audition, Paris (IDA; France), providing the basis for evaluating the comparability of measurements across centres and countries.

In this paper, the PRESAGE test battery is introduced in detail, the parameters of interest for each assessment are described, and data from the project’s control group are reported, providing a baseline for interpreting variability across participants. The control group comprises participants aged 40 years and older, with a PTA lower than the age-dependent median according to ISO 7029 (2017) in the better ear. Therefore, this dataset also establishes a foundation for future analyses of age-matched comparisons of test battery outcomes against the control group, multidimensional auditory profiling, and genotype-phenotype correlations.

### Composition of test battery

A distinctive feature of the PRESAGE test battery is its systematic assessment of suprathreshold deficits in combination with standard audibility, physiological, and vestibular measures, extending beyond previous test batteries and enabling a truly multidimensional characterisation of hearing function. Although genetic analyses are not yet included in the present publication, future work will combine this multidimensional phenotyping with genetic data, offering unprecedented insights into the contribution of genetic factors to individual patterns of hearing loss. To our knowledge, such comprehensive reference data across functional, physiological, vestibular, and genetic domains is unparalleled in hearing research. This approach holds considerable potential for advancing individualised diagnostic protocols and guiding targeted interventions in audiology.

Specifically, the audiological functional tests comprise adaptive categorical loudness scaling (ACALOS; Brand & Hohmann, 2002; Oetting et al., 2014), which captures individual loudness perception and binaural loudness summation; speech recognition in noise assessed with the Matrix sentence test in German and French (Kollmeier et al., 2015), a broadband suprathreshold measure with high ecological validity due to the relevance of speech in noise perception in daily communication; and tone-in-noise detection (TIN; Hülsmeier & Kollmeier, 2022), which provides frequency-specific insights into suprathreshold processing deficits, particularly related to intensity resolution. These tests therefore represent complementary suprathreshold components of hearing loss. For both the Matrix test and TIN, audible noise levels were chosen to ensure that the tests effectively characterise suprathreshold deficits rather than audibility limitations.

The audiological physiological tests comprise electrocochleography (ECochG, Oku & Hasegewa, 1997; Konrad-Martin et al., 2012), which measures cochlear and auditory nerve potentials to assess peripheral auditory function, and otoacoustic emissions (OAE, Bertoli & Probst, 1997; Kemp, 2002; Hoth et al., 2010; Vaden et al., 2018), which characterise outer hair cell function responsible for cochlear amplification and dynamic compression. Furthermore, the middle-ear muscle reflex (MEMR) is included, which examines the integrity of the middle-ear reflex pathway and associated brainstem circuits (Silman & Gelfand, 1981; Guest et al., 2019). Together, these measures provide a complementary physiological assessment that can be related to suprathreshold functional deficits.

The vestibular physiological tests include the caloric test (Barin, 2008; Shepard & Jacobson, 2016), the video head impulse test (vHIT; Murnane et al., 2014; Halmagyi et al., 2017), and vestibular evoked myogenic potentials (VEMP; Colebatch et al., 1994; Todd et al., 2007; Rosengren et al., 2010). While the former two tests characterise the function of the semicircular canals at different head velocities, the latter evaluate otolith function at the sacculus and utriculus. In combination, these tests provide a comprehensive characterisation of vestibular function.

### Objectives

This paper aims to provide a multidimensional characterisation of both functional and physiological aspects of hearing and balance functions using a comprehensive audiological-vestibular test battery, including comprehensive information about test methods and conditions to allow full reproducibility. As a first step, univariate relationships between the included measures and age and PTA are assessed as a baseline for independent age dependency and contributions beyond audibility. Specifically, the study seeks to:

1. Develop a comprehensive test battery of audiological-functional, audiological-physiological, and vestibular-physiological tests that allows for suprathreshold and cross-domain characterisation of both hearing and balance functions.
2. Evaluate across-site comparability of data collected in Oldenburg and Paris, accounting for differences in equipment or protocols, and establish a harmonised framework for interpreting results across centres.
3. Examine how functional and physiological measures vary with age and PTA in the project’s control group of German and French participants with age-normal hearing thresholds.
4. Provide reference data for the control group to support pooled analyses, explore genotype-phenotype relationships, and investigate correlations across functional, physiological, and vestibular measures to lay the foundation for future multidimensional analyses.

## Materials and Methods

### Participants

Fifty-five adults aged 40 years and older took part in the study (mean age 64.9 ± 9.8, range: 40 - 89 years; 36 females). Of these, thirty-nine were recruited and tested at the University of Oldenburg (UOL), Germany (mean age 64.8 ± 10.1, range: 40 - 89 years; 27 females), and sixteen at the Institut de l’Audition (IDA), Paris, France (mean age 65.0 ± 9.4, range: 42 - 78 years; 9 females). Participants were required to have age-appropriate symmetric normal hearing, defined as better-ear pure-tone average (PTA) across the frequencies 0.5, 1, 2, and 4 kHz better than the age- and sex- specific median threshold specified in ISO 7029 (2017) and less than 10 dB PTA difference between sides. Additional inclusion criteria were a normal middle-ear status (assessed via tympanometry and otoscopic inspection, no conductive hearing loss according to bone conduction hearing thresholds), no self-reported hearing-related problems or comorbidities such as diagnosed diabetes II, cancer, neurological or cognitive disorder, or diagnosed noise-induced hearing loss. Although three participants (UOL: 2; IDA: 1) reported type II diabetes, their inclusion was retained due to the absence of hearing-related complaints or comorbidities relevant to the study. All participants reported being native speakers of the test language–German at UOL and French at IDA. UOL participants were recruited from the Hörzentrum Oldenburg participant registry, and IDA participants were recruited through study information posters and flyers inviting participants to contact the research team. All participants gave written informed consent as approved by the pre-medical Ethics Committee of the Universität Oldenburg (2022-074) and the Comité de Protection des Personnes (CPP; Committee for the protection of persons) Sud Ouest et Outre-mer 1 (2021-062). All participants received monetary compensation for their participation.

### Procedure

Testing comprised an inclusion part and a follow-up part: (1) a screening session including a questionnaire covering general health (e.g., diagnosed diabetes II, cognitive or neurological disorders, neck pain, visual impairment, and risk of falling) and hearing-related aspects (e.g., diagnosed hearing loss, duration, degree, known cause, family history, history of middle ear infections, and various aspects concerning occupational and recreational noise exposure). This was followed by an ear canal examination with cerumen removal if the tympanic membrane was not visible, middle-ear function assessment (tympanometry), pure-tone audiometry (air conduction, AC at 0.125, 0.25, 0.5, 1, 2, 3, 4, 6, and 8 kHz; bone conduction, BC at octave frequencies 0.5 – 4 kHz); (2) follow-up sessions comprising audiological-functional, audiological-physiological, and vestibular-physiological assessments from the test battery. The follow-up sessions lasted about two hours each, and participants completed one or several sessions either in a single visit or across separate visits, depending on their preference and availability. In case of multiple visits, each started with otoscopic inspection and tympanometry to reconfirm unobstructed ear canals and middle-ear status (e.g., no congestion or pressure changes).

At UOL, the screening and vestibular tests were conducted at Evangelisches Krankenhaus, Oldenburg, Germany, and the psychoacoustic and audiological-physiological tests at the university’s Medical Physics laboratory. Audiological measures at the university laboratory were always performed in a sound-attenuated and electromagnetically shielded booth. At the hospital, assessments were conducted either in a sound-treated room or, when unavailable, in a quiet office. At IDA, all follow-up sessions took place in sound-attenuated and electromagnetically shielded booths at the Centre for Research and Innovation in Human Audiology (CERIAH), Institut Pasteur, Paris, France. Inclusion visits were conducted at the CERIAH and at one Audition Santé (Sonova Audiological Care France) hearing centre in Paris (by author AC).

### Audiological functional measures

#### Adaptive categorical loudness scaling

The Adaptive Categorical Loudness Scaling test (ACALOS; Brand & Hohmann, 2002) is a standardised measure of loudness perception (ISO 16832, 2006) and has been shown to be effective in hearing aid fitting (Oetting et al., 2018). Participants self-rated the loudness of a stimulus on an 11-point scale, corresponding to categorical units (CU, 0 – 50), ranging from ‘not heard’ to ‘too loud’. The procedure consisted of two measurement phases. In the first phase, the individual dynamic range was estimated using an adaptive, interleaved loudness scaling procedure, continuing until the responses ‘not heard’ and ‘too loud’ were obtained. In the second phase, stimuli were presented in a randomised order within the individually determined dynamic range. The maximum presentation level was 100 dB SPL, and each stimulus had a duration of 1 second, shaped with 50 ms von Hann window ramps at onset and offset. Broadband noise (IFNoise; Holube et al., 2008) was presented under both binaural and monaural configurations in a counterbalanced order across participants.

Measurements at both centres were performed using the Oldenburg Measurement Applications software (OMA; Hörzentrum Oldenburg gGmbH), a high-power external sound card (EarBox 3.0; AURITEC GmbH, Hamburg, Germany), and circumaural headphones – Sennheiser HDA 200 (Wedemark, Germany; UOL) or RadioEar DD 450 (New Eagle, PA, USA; IDA) headphones. Headphones were free-field equalised according to ISO 389-8:2004 and ISO 389-1:2017. Calibration of the noise was carried out in OMA using a Brüel & Kjær (B&K; Sound and Vibration Measurements A/S, Nærum, Denmark) 2610 amplifier, 4153 artificial ear, 4134 ½-inch microphone, 2669 microphone preamplifier, and a 4192 microphone capsule.

Individual loudness curves were fitted in MATLAB using the BX method (Oetting et al., 2014) via the function *fit_loudness_function,* available in the AFC toolbox^1^ (version 1.4.1; Ewert, 2013). The BX method is a modification of the approach by Brand and Hohmann (2002) that minimises fitting error in stimulus level (i.e., x-axis) direction and reduces the influence of extreme outliers (>40 dB), thereby improving the robustness of the fit. The resulting fit is described by three parameters: m_low_ and m_high_, representing the slope below and above 25 CU, respectively, and L_cut_, the intersection point of the two lines at 25 CU. Four outcome parameters were defined for analysis: estimated levels (dB SPL) at ratings of 15 CU (‘soft’, L15) and 35 CU (‘loud’, L35), the dynamic range (DR) calculated as a difference between L35 and L15, and binaural loudness summation (BLS). BLS was defined as the difference between the estimated level in the binaural and monaural configuration (average of the left and the right ears) as described in van Beurden et al. (2018).

ACALOS testing preceded all other suprathreshold functional measures and was used to establish a reference level of ‘medium’ loudness (25 CU) at the listener’s better ear to ensure that the noise was audible in Matrix test and TIN, and hence both tests were effectively suprathreshold. If the measured level deviated by more than 5 dB from the IFNoise normative level for normal-hearing adults (70.2 dB SPL; see Table 3 in Oetting et al., 2018), the noise level in the subsequent measures was increased accordingly, up to a maximum of 20 dB. This rule was introduced after the first few participants and applied consistently thereafter. As the participants had no hearing impairment and were instructed to report if noise was inaudible, it is reasonable to assume that all subsequent measures were performed in noise as intended.

#### Tone-in-noise

The tone-in-noise (TIN) test was included to assess frequency-specific tone detection thresholds in noise. The TIN threshold reflects the listener’s ability to detect tones in noise beyond audibility. It captures individual variability in auditory resolution and has been linked to speech recognition performance in noise, even when hearing sensitivity is within normal limits (Schädler et al., 2020; Hülsmeier & Kollmeier, 2022). Absolute detection thresholds were measured monaurally for the participants’ better ear according to PTA. TIN was assessed using the graded response bracketing (GRaBr) adaptive procedure, which has been shown to improve robustness against listener inattention and enhance threshold estimation (Xu et al., 2024). Listeners indicated whether they heard two tones, one tone, or none, with a louder cue tone included to guide and control for attention. The level difference between probe and cue tones was adaptively reduced, starting at 10 dB, using a staircase procedure similar to that of the probe tone, making them gradually closer in level to prevent reliance on relative loudness cues. To control for response reliability, 20% of the trials were catch trials in which the cue tone was muted. Thresholds were measured at 0.5 and 2 kHz, presented in a two-octave-wide white noise centred on the target frequency and with a duration of 1600 ms. To avoid off-frequency tone detection, two flanking noises were added to the masker: an f-flanking noise (blue noise, with a power spectral density increasing by 3 dB per octave and low-pass filtered at the lower cutoff frequency of the bandpass masker), and a 1/f-flanking noise (pink noise, with a power spectral density decreasing by 3 dB per octave and high-pass filtered at the upper cutoff frequency of the bandpass masker). The spectral density levels of the flanking noises were set 6 dB below the level of the two-octave-wide white noise at their respective cutoff frequencies. The spectral noise level was individually set between 30 and 50 dB SPL, based on the listener’s medium loudness perception (see ACALOS). The procedure began with a 90 dB SPL probe tone level and 8 dB step size, decreasing to 2 dB after three reversals. Thresholds were calculated as the median of the final 10 of 14 reversals. Testing began with a 1 kHz practice run, which was excluded from analysis. Absolute thresholds were converted to SNRs relative to the masker level within the probe tone’s equivalent rectangular bandwidth (ERB; Moore and Glasberg, 1983), accounting for individually adjusted noise levels and masker energy at the probe tone.

Custom Octave scripts controlled the test, adapted from Schädler et al. (2020). Stimuli were presented using the same headphones and sound card, and calibration of the noise was carried out with the same equipment as described for ACALOS, with the same calibration script as in Schädler et al. (2020).

#### Matrix sentence test

Speech recognition in noise was assessed using the matrix sentence test (Matrix). It was selected for its high diagnostic sensitivity to sensorineural hearing loss and its validated cross-language comparability, including the German and French versions (Kollmeier et al., 2015). Speech material consists of 5-word unpredictable sentences with a fixed grammatical structure. Speech recognition thresholds (SRTs), defined as the signal-to-noise ratio (SNR) at which 50% of the words are correctly recognised, were measured using an adaptive procedure (A1; Brand & Kollmeier, 2002) with constant noise level between 65 and 85 dB SPL based on the listener’s medium loudness perception (see ACALOS), and word scoring. Each test list consisted of 20 sentences. Language-specific versions were used depending on the centre: German (Wagener et al., 1999a, b, c) at UOL and French (Jansen et al., 2012) at IDA. Two types of noise were used, each targeting different aspects of auditory processing: test-specific (and language-specific) stationary speech-shaped noise (SSN), reflecting primarily energetic masking, and speech-modulated noise (ICRA5-250, Wagener et al., 2006), targeting spectro-temporal processing mechanisms such as dip listening and modulation detection. ICRA5-250 was chosen as fluctuating noise in both languages although a female voice was used in the French Matrix test. SRTs in ICRA5-250 and ICRA4-250, exhibiting a male and female speech spectrum, respectively, were shown to be comparable (Wagener et al., 2006; Hochmuth et al., 2015). Two training lists were presented to mitigate learning effects. Testing was conducted monaurally for the better ear for both SSN and ICRA5-250 noise. The order of the noise conditions and the choice of test lists were randomised across listeners. The same setup and calibration procedure as for ACALOS was applied.

To enable centre-independent analysis, SRTs were expressed as reference-normalised SRTs (SRT_norm_), calculated by subtracting the language- and noise-specific normative SRT from the measured value. Normative values for young normal-hearing listeners (monaural presentation at better-ear, word scoring) were –7.1 dB SNR for SSN, with a psychometric slope of 17.1 %/dB (Wagener et al., 1999a, b, c), and –19.3 dB SNR for ICRA5-250 (Hochmuth et al., 2015) for the German Matrix test, and –6.0 dB SNR for SSN, with a slope of 14.0 %/dB, and –16.1 dB SNR for ICRA4-250 for the French Matrix test (Jansen et al., 2012). The resulting SRT_norm_ values therefore reflect the deviation from young normal-hearing performance on a common scale, with SRT_norm_ = 0 dB indicating normative performance.

### Audiological physiological measures

#### Middle-ear muscle reflex

Middle-ear muscle reflex (MEMR) thresholds were measured using an immittance meter with a probe inserted into the external auditory canal with an airtight seal emitting a 226 Hz pure tone. MEMR thresholds were elicited ipsilaterally at 500 Hz, 1000 Hz, and 2000 Hz. Stimulus intensity increased from 80 to 100 dB nHL in 5 dB steps until a reproducible reflex response was obtained or the maximum level was reached (i.e., 100 dB nHL). Reflex threshold was defined as the lowest intensity eliciting a measurable and repeatable contraction. Participants were seated in an anechoic or quiet room and instructed to maintain a relaxed posture, with no active response required. Measurements in both centres were performed with the Interacoustics Titan IMP440 Impedance System (Interacoustics A/S, Middelfart, Denmark). A reflex was considered as triggered by the software if a variation of the volume of the ear canal of at least 0.02 ml was detected.

#### Otoacoustic emissions

Transient evoked otoacoustic emissions (TEOAEs) and distortion product otoacoustic emissions (DPOAEs) were recorded in a sound-attenuated room using an ear probe equipped with an acoustic transducer and a high-sensitivity microphone, connected to the Elios device (ECHODIA, France). The probe was fitted with an ear tip whose diameter was adapted to each participant’s ear canal morphology to ensure optimal sealing and prevent acoustic leaks. Probe stability was maintained throughout the measurement and monitored visually to ensure reliable recordings. The calibration of the device was regularly verified with the test cavity provided by the manufacturer.

TEOAEs were elicited using click stimuli at 85 dB peak SPL, covering a frequency range from 1 to 5 kHz. TEOAE presence at each frequency was determined by the software according to combined SNR and response reproducibility criteria (SNR ≥ 9 dB and reproducibility ≥ 50%, SNR ≥ 6 dB and reproducibility ≥ 60%, or SNR ≥ 3 dB and reproducibility ≥ 75%). DPOAEs were recorded using two simultaneous primary tones (f1, f2; f2/f1 ≈ 1.22), eliciting a distortion product at 2f1–f2 that reflects outer hair cell integrity in the cochlear region tuned to f2 (Kemp, 1979; Probst et al., 1987). Stimulus levels were L1 = 65 dB SPL and L2 = 60 dB SPL, with both intensities increased to 68 dB SPL if a response was detected at less than two of the measured frequencies. Frequency-specific responses were recorded across the f2 range (1 to 4 kHz) to generate DP-grams. At IDA, DPOAEs were additionally recorded at further f2 frequencies, but only the frequencies common to both centres (1, 2, and 4 kHz) were used for the analysis and for determining whether intensity levels were increased. DPOAE presence per frequency was defined as an SNR ≥ 6 dB and a DP level of ≥ -6 dB.

#### Electrocochleography

Electrocochleography (ECochG) recordings were performed using the Elios device via the Echosoft software (ECHODIA, France). Participants were comfortably seated in a reclining armchair and instructed to minimise movements to reduce muscular artifacts. Two intra-auricular Gold Eartip Electrode (TIPtrode ER3-26A) electrodes delivered calibrated acoustic stimuli directly into both ear canals, while electrical responses were recorded via two surface electrodes placed on the forehead, with the midline electrode serving as ground and the lateral electrode as reference. The stimulation protocol consisted of 100 µs alternating-polarity clicks, presented at a rate of 11 stimuli/s. A 2 ms pause was included before each stimulus to allow a better baseline analysis. For each condition, 1,000 responses were averaged to improve the signal-to-noise ratio, with automatic rejection of trials exceeding 20 µV in amplitude (rejection window starting at -2 ms). Signals were acquired at 32 kHz and band-pass filtered between 100 Hz and 4 kHz. Clicks were presented at intensities of 85, 95, 105, and 115 dB peak SPL (corresponding to 60, 70, 80, and 90 dB nHL, respectively).

Analysis focused on the presence and latency of the characteristic waves I, III, and V (Boston et al., 1985). Latencies were measured as the time interval between stimulus onset at the eardrum (0 ms) and the peak of each detected wave. The presence or absence of each wave was determined for each stimulus intensity. The waves were manually labelled within the software by the authors SK and GG. To assess inter-rater consistency, both authors independently labelled the recordings for UOL data (39 of 55 participants), and the resulting wave latencies and presence/absence decisions were compared. Agreement within 0.15 ms was found for 54.4 % of all waves. For the remaining waves, a consistent compromise between presence and absence of the waves was identified, resulting in a balanced choice of labels from the two raters.

### Vestibular physiological measures

#### Video head impulse test

The response of each semicircular canal (SCC) to high-frequency stimulation can be measured individually with the video head impulse test (vHIT; MacDougall et al., 2013a; MacDougall et al., 2013b; McGarvie et al., 2015). At IDA, vestibulo-ocular reflex (VOR) measurements were performed using a remote camera system (Synapsys-Inventis®, France). This system utilises binocular eye movement recording via an infrared video camera (100 Hz sampling rate) mounted on a floor stand at 90 cm distance. Participants were seated on a chair with their eyes fixated on a stationary visual target located 1 to 1.3 m straight ahead. Prior to testing, calibration was performed for each participant using the system’s built-in calibration process. The camera placement was adjusted to align horizontally and vertically with the participant’s eyes, and the camera position was fine-tuned within the video frame to ensure optimal pupil detection. For optimal recording, subjects were instructed to keep their neck relaxed and teeth clenched (to reduce movement artifacts) and to keep their eyes wide open without blinking. To reduce stiffness artefacts, participants were asked to keep their arms resting alongside the body.

At UOL, testing followed the same seating and fixation setup, but VOR measurements were carried out using the ICS Impulse USB system (Otometrics, Natus Medical, Denmark) with right monocular infrared video goggles (sampling rate 250 Hz) tightly fixed on the head with an elastic band. A single-use face cushion improved comfort and reduced slippage, and the USB cable was clipped to the garment to minimise artefacts during movement. Fixation alignment was verified by matching the two laser beams from the goggles to the target before performing the automatic calibration, helping the participant to keep the head steady while following a single moving dot.

For both systems, calibration and pupil-tracking optimisation were conducted prior to testing, following the manufacturers’ recommended procedures to ensure correct gaze alignment, minimise artefacts, and optimise tracking quality. All procedures adhered to the manufacturer’s guidelines and established clinical protocols to assess VOR function across all six semicircular canals (Murnane et al., 2014). The impulses were tested in six distinct directions, stimulating the right and left horizontal (lateral) canals, as well as the left anterior–right posterior (LARP) and right anterior–left posterior (RALP) pairs for the vertical canals. For certain vertical canal impulses, the head was pre-rotated 35°–45° in the yaw plane. The examiner administered head impulses while standing behind the participant, placing their hands on the forehead (Patterson et al., 2015). Head impulses were passive, sudden, and unpredictable to avoid voluntary prediction or compensation (Della Santina et al., 2002). The amplitude of head impulses was low, about 10°–20°, with high velocity and high acceleration. Impulses were delivered until at least five to seven valid responses were obtained per canal, ensuring that they met hardware and software criteria for velocity, acceleration, trajectory, and pupil detection.

The VOR gain was calculated by the software as the ratio of mean eye velocity (°/s) to mean head velocity (°/s), measured as the area under the velocity curves within a selected time interval during the impulses. To assess interaural differences in canal responses, an asymmetry ratio (AR, expressed in %) was calculated for each canal pair using Equation 1 (Striteska et al., 2025). The same equation was used for data collected in both centres, to ensure comparable estimates despite differences between device-specific calculation methods. Note that the main difference between the used devices was the binocular (Synapsys) versus monocular (Otometrics) recording, which introduces a systematic asymmetry bias for measurements with the monocular system.

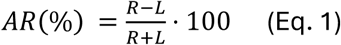

where R and L denote the mean VOR gains for the right- and leftward head impulses, respectively. Positive AR values therefore indicate higher VOR gain on the right side, and negative values higher gain on the left side. To enable direct comparison of asymmetry direction across participants relative to their respective better ear, we inverted the AR sign for all participants with left better ear according to PTA. The rationale behind this was to assess whether the vestibular asymmetry corresponds to the PTA asymmetry direction. Consequently, positive AR indicates a stronger response on the PTA better ear side, whereas a negative AR indicates a stronger response on the PTA worse-ear side.

#### Vestibular-Evoked Myogenic Potentials

Vestibular-evoked myogenic potentials assess the function of saccular and utricular otolithic receptors. Cervical vestibular-evoked myogenic potentials (cVEMPs) were measured to evaluate saccular and inferior vestibular nerve function, and ocular vestibular-evoked myogenic potentials (oVEMPs) to evaluate utricular and superior vestibular nerve function (Colebatch et al., 1992; 1994). Testing was performed bilaterally under standardised conditions using a Neuro-Audio system (Neurosoft, Ivanovo, Russia) at IDA and an Eclipse system (Interacoustics A/S, Middelfart, Denmark) at UOL.

VEMPs were elicited using 750 Hz tone bursts (2 ms rise/fall, 0 ms plateau) delivered via insert earphones (Radioear IP30). Stimulation rates were set to 5 Hz. Testing began at 95 dB nHL, and the level was increased in 5 dB steps to determine the response threshold. For analysis, recordings obtained 5 dB above threshold were used to ensure reliable response identification. The maximum stimulus level was 110 dB nHL, when tolerated. Each recording consisted of 50–100 stimuli per ear, with a minimum of 2–4 repetitions to ensure response reproducibility. For both cVEMPs and oVEMPs, biphasic waveforms (P1 and N1) were accepted if reproducible across at least two successive repetitions at the same stimulus level.

For cVEMP, participants were positioned supine with the torso inclined at 30°. Active (recording) electrodes were placed over the upper half of the ipsilateral sternocleidomastoid (SCM) muscle, with the reference electrodes on the sternoclavicular junction, and the ground electrode on the forehead (van Stiphout et al., 2022). Participants were instructed to turn their head contralateral to the stimulated ear to ensure sufficient SCM activation, targeting a minimum tonic electromyographic (EMG) activity (Curthoys, 2010; Colebatch and Rothwell, 2004). EMG signals were amplified and bandpass filtered between 1 and 1,000 Hz (IDA) and between 10 and 1500 Hz (UOL) as recommended by the respective device manufacturer. Response parameters included peak latencies and amplitudes of characteristic biphasic waveforms at P1 (positive peak) and N1 (negative peak) from the ipsilateral SCM.

For oVEMP, participants were seated supine (IDA) or upright (UOL) with their head supported and instructed to fix their gaze upwards (20°–30°) on a target placed 1 to 2 m away to optimise eyeball elevation and maximise response amplitude. Surface active electrodes were placed 1–3 mm below each lower eyelid (Ochi et al., 2003). At IDA, reference electrodes were placed on the forehead (at the nasion) (van Stiphout et al., 2022) and the ground electrode on the sternoclavicular junction, whereas at UOL, reference electrodes were aligned directly below the active electrodes and the ground electrode was placed on the forehead. oVEMPs were recorded as short-latency, biphasic EMG responses at N1 (negative peak) and P1 (positive peak), typically from the contralateral inferior oblique muscle. Recorded waveforms were accepted if reproducible and within reference latency and amplitude ranges. EMG signals were amplified and bandpass filtered between 30 and 2,000 Hz (IDA) and between 1 and 1500 Hz (UOL).

Raw VEMP peak-to-peak amplitudes (absolute difference between P1 and N1) were normalised by background EMG activity to obtain comparable normalised amplitudes across participants. Asymmetry was calculated as for vHIT (Equation 1), and was expressed relative to the participant’s PTA better ear.

#### Caloric test

Caloric testing was conducted to evaluate the function and symmetry of the vestibulo-ocular reflex (VOR), with specific focus on the function of the horizontal (lateral) semicircular canals under low-frequency stimulation (Janky and Rodriguez, 2018). Participants were informed about possible post-test dizziness and advised against driving immediately afterward due to potential residual dizziness. Testing was conducted as the final procedure in the vestibular battery. Participants were positioned supine with their head elevated approximately 30° above the horizontal plane, aligning the lateral semicircular canals vertically to optimise endolymphatic response during thermal stimulation. Testing was performed in a darkened room with vision denied to minimise visual fixation and suppress optokinetic responses.

Caloric testing was performed using a validated water irrigation device (Variotherm Plus, ATMOS), and eye movements were recorded with infrared video nystagmography (VNG) systems (IDA: Ulmer VNG, Synapsys, France; sampling rate: ∼100 Hz; UOL: ICS Impulse USB system, Otometrics, Natus Medical, Denmark with monocular high-speed infrared video goggles, sampling rate 173 Hz). Data were acquired and analysed via the Ulmer VNG software and the OTOsuite vestibular software (Otometrics), respectively. Spontaneous nystagmus was assessed and corrected before irrigation commenced. Standard bithermal water irrigation was delivered according to the Fitzgerald-Hallpike protocol, using cold (30°C) and warm (44°C) water stimuli per ear. Irrigation volumes ranged from 20 mL to 300 mL, administered over a duration of 30 seconds, adjusted according to participant tolerance. Volumes and durations were kept consistent for both ears to maintain comparability across sides. Between successive irrigations, a five-minute interval was used to ensure that the nystagmus from the preceding irrigation had fully resolved. In cases of intolerance (e.g., pronounced vertigo, vegetative reactions, nausea, or excessive discomfort), stimulus duration was proportionally reduced (e.g., to 15 seconds) or the measurement was stopped after irrigation with one temperature (monothermal stimulation; occurring in one IDA participant).

Oculomotor responses were recorded continuously, monitoring eye movements monocularly throughout irrigation to capture both peak and sustained nystagmus responses. The primary quantitative parameter was the maximum slow-phase velocity (SPV) of caloric-induced nystagmus, measured in degrees per second (°/s). Key outcome measures included the asymmetry metrics unilateral weakness (UW) and directional preponderance (DP) (McCaslin et al., 2014), calculated according to the Jongkees or Phillipszoon-Jongkees formulas (Jongkees et al., 1962; Equations 2 and 3). For comparability across devices and centres, absolute SPVs were used, accounting for different output formats of the devices. Unilateral weakness quantifies interaural side differences, with positive/negative values describing a reduced responsiveness of the left/right ear, respectively. Directional preponderance quantifies differences between responses of right- and left-beating nystagmus, taking into account that the nystagmus is beating right for warm-right (RW) and cold-left (LC) stimulation, and beating left for warm-left (LW) and cold-right (RC) stimulation. Hence, positive DP indicates a right-beating nystagmus. In our analysis, both UW and DP were expressed relative to the participant’s PTA better ear by multiplying all responses for participants with left better ear with -1. This leads to the interpretation of positive UW (Equation 2) as unilateral weakness of the worse ear, and positive DP (Equation 3) as nystagmus beating in the direction of the better ear. For the participant with cold monothermal stimulation, the monothermal caloric asymmetry (MCA) was calculated as alternative asymmetry metric, but excluded from subsequent analysis due to inadequate diagnostic accuracy of cold MCA (Lightfoot et al., 2009). Complete absence of caloric response would be interpreted as vestibular paralysis.

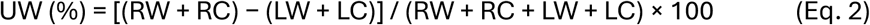

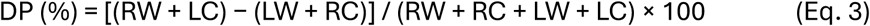

### Statistical analysis

#### Data preprocessing

Data analysis was performed with R version 4.3.2 (R Core Team, 2024). The full reproducible code is available on Zenodo. Raw data from all tests and participants was extracted by either automatically reading from export files in XML or XLSX format if available, or manually documented in XLSX tables based on PDF output files. All data chosen for this paper was combined in one XLSX file, integrating French and German data. The better ear was estimated by calculating the minimum PTA across ears. Missing data were assessed with respect to the data required for the analysis in this paper. Of the 55 included participants, three did not complete TIN due to time constraints, five wished not to perform the caloric test, and one data point for binaural ACALOS was excluded from analyses due to a technical error.

#### Centre effect

The comparability and compatibility of French (IDA) and German (UOL) data were investigated as a prerequisite for combining data from both centres in the following analysis. The centre effect was assessed by a generalised linear model (GLM; Jiang and Nguyen, 2021), as defined in Equation 4, with 𝑐𝑒𝑛𝑡𝑟𝑒 as factor (IDA as reference level), and 𝑃𝑇𝐴 and 𝑎𝑔𝑒 as covariates to account for PTA- and age-dependency, reflecting the variables used in the subsequent analyses. The centre effect for the respective variable 𝑣𝑎𝑟_𝑖_ was defined as present if the coefficient 𝛽_1_ was significant (p < 0.05), indicating a significant difference between French and German data.

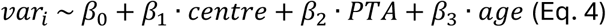

To allow for a fair comparison between centres, the centre effect calculation was performed on a matched subset of IDA (n=16) and UOL (n=39) participants. Participants were matched by PTA and age based on the nearest neighbour matching procedure performed with the R package matchit (Ho et al., 2011). The Mahalanobis distance was used as distance measure, as it provides a robust (deterministic) calculation for small sample sizes (Li et al., 2019). A one-to-one matching of French and German participants was performed, resulting in a matched data set of n=16 participants per centre. The successful matching was assessed by evaluating the balance of the two data sets with respect to the adjusted estimated mean difference (β_1_), standard error (SE), p-value, and effect size (Cohen’s d). As additional pre-analysis for the GLM, Levene’s test was performed to assess homogeneity of variance for all variables 𝑣𝑎𝑟_𝑖_ (Fox and Weisberg, 2019). For the Matrix test, the centre effect was assessed on the language-specific data (SRT) as well as on the reference-normalised data (SRT_norm_).

#### Age and PTA effects

The effects of age and better-ear PTA on the selected outcome variables were assessed using Spearman’s rank-order correlations (stats package; R Core Team, 2024) with age and PTA treated as continuous predictors. Age and PTA were analysed independently to identify any existing effect as a first step. Related to the choice of the control group covering only a small PTA range, no or only small PTA effects were expected. Future analyses on larger PTA ranges could control for interactions between PTA and age to take into account known correlations. Scatterplots were used for visualisation, with added linear regression lines to guide interpretation. To aid visual interpretation, participants were grouped into 10-year age bins from 40 to 90 years and colour-coded in the figures, with UOL participants indicated by a black outline. Spearman’s correlation coefficient (ρ), associated p-value, and number of observations (n) are reported in each plot, with the number of UOL participants shown in parentheses.

#### Effect of loudness category on binaural summation

To evaluate whether binaural loudness summation differs between loudness categories (L15 and L35), a linear mixed-effects model (LME) was fitted using the *lmer* function from the lme4 package (Bates et al., 2015), with loudness categories (L15/L35), and PTA as fixed effects, and participant as random intercept. Model assumptions of normality and homogeneity of variance were met, as confirmed by Shapiro-Wilk and Levene’s tests, respectively. Model comparisons were performed using maximum likelihood estimation and the final model was refitted using restricted maximum likelihood (REML) for reporting. Degrees of freedom were approximated using the Kenward-Roger method.

## Results

### Overview

All 55 participants showed a symmetric PTA with a median interaural difference of 2.5 dB (IQR [1.25, 4.00] dB, max. 8 dB). For subsequent analyses, data for the better ear according to PTA are presented for all monaural tests and variables, whereas data from both ears are reported when appropriate (e.g., binaural ACALOS condition or for asymmetry measures in vestibular tests).

Figure 1A shows better-ear PTA over age for all IDA and UOL participants. All PTA values are below the age-specific median thresholds of ISO 7029 (2017) for female (blue line) as defined in the inclusion criteria, within a 5 dB error margin. Participants are depicted in their respective age-group colour, with unmatched participants represented by reduced opacity. Figure 1B depicts the corresponding distributions of age and PTA for all participants by centre. While the age distributions are similar between centres, many UOL participants exhibit lower PTA values, often below 10 dB HL. After matching, PTA distributions are more similar, while the age distributions show slightly greater differences (Figure 1C), resulting in balanced groups suitable for assessing the centre effect. The matched pairs, as visualised in Figure 1A with the connecting grey lines, are generally close in the PTA-age space, except for a few points with greater distances at higher ages.

**Figure 1.**
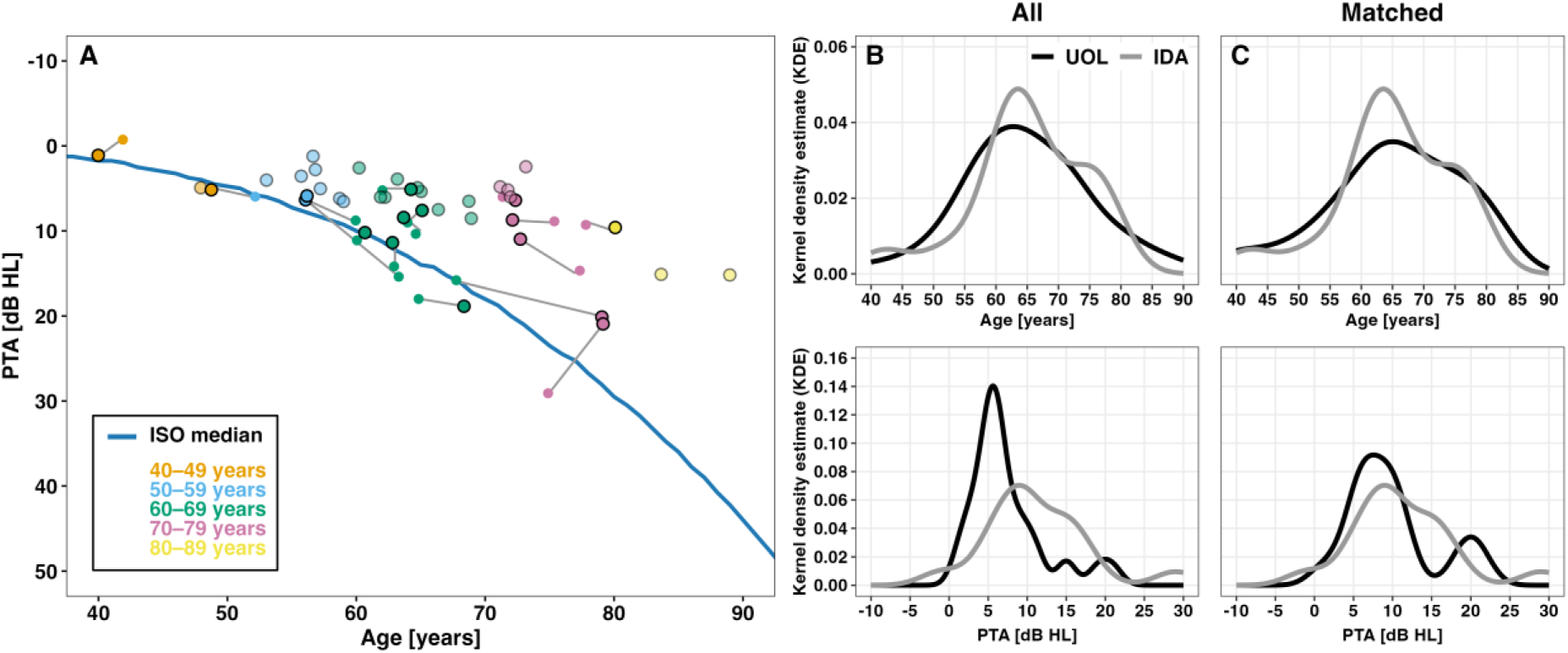
Better-ear PTA as a function of age and age/PTA distributions before and after centre matching. (A) Better-ear PTA values are shown across all participants from IDA and UOL in relation to the age-specific normative median thresholds for females (ISO 7029, 2017; blue line). UOL participants are marked with black-outlined circles, while IDA participants are shown without outline. Matched pairs are connected by grey lines. Unmatched participants are displayed with reduced opacity. Colours indicate age groups by decades. Distributions of age and better-ear PTA are shown for the full sample (B) and for the subset of matched participants (C), plotted separately by centre (UOL in black, IDA in grey).

These are relatively rare and occur due to the age dependency of the ISO 7029 (2017) norm, according to which higher PTA values are below the median in older individuals.

### Centre effect

The statistical analysis of the centre effect was carried out on a subset of 16 matched participants per centre. GLM and Levene’s test results are summarised in Table A1. The results directly support combining data from both centres for subsequent analyses for most of the considered variables. However, a significant centre effect was found for the ACALOS level variables (L15: β_1_=-5.14 dB, p<0.01; L35: β_1_=-5.08, p<0.05), both language-specific Matrix test conditions (SSN: β_1_=1.49 dB, p<0.01; ICRA5-250: β_1_=3.18 dB, p<0.001), and VEMP normalised peak amplitudes (cVEMP: β_1_=0.35 µV, p<0.05; oVEMP: β_1_=0.56 µV, p<0.001). Centre differences were also found for several single variables, including caloric test SPV for cold irrigation (β_1_=7.78 °/s, p<0.05) and vHIT lateral canal gain (β_1_=-0.12, p<0.01). For the Matrix test, the centre effect observed in the language-specific SRTs was expected due to the different normative values of the French and German test versions. Normalising the SRTs to the test-specific references (SRT_norm_; see Methods) eliminated the centre effect in both noise types, indicating comparable performance across centres. Consequently, the combined SRT_norm_ values were used in the following analyses. ACALOS estimated levels, although significantly different between centres, were combined for subsequent analysis, as the observed difference (∼5 dB) falls within the expected inter-individual variability of this measure (Oetting et al., 2014). The centre differences in the VEMPs’ normalised peak amplitudes are likely due to the different devices used at IDA and UOL. For tests and variables that showed a small but significant centre effect in the GLM, their interpretation and comparability is assessed below together with information on the test procedure, equipment, and visual inspection of the data in the respective sections. Given the low number of participants per centre (n=16) included in the centre effect analysis, it is likely for a significant centre effect to be found. This is due to the fact that PTA and age values do not always closely align. Importantly, variability across participants is expected for the variables assessed in the proposed test battery, as their ability to capture information beyond PTA is central to the objectives of the present study. Overall, the data from IDA and UOL appear to be comparable. Based on the investigations on the matched group, the following analyses were conducted on all data from IDA (n=16) and UOL (n=39).

### Age and audibility effects

To examine the influence of age and hearing sensitivity on performance, the relationships between the selected outcome variables and age and PTA, respectively, were analysed using Spearman’s rank-order correlations for all tests. Table 1 summarises the outcome variables across all participants, or by centre (Matrix test), reported as median with interquartile range (IQR), together with linear regression coefficients (β_0_, β_1_), Spearman’s rank correlation (ρ), and corresponding p-values. Table A2 provides median and interquartile ranges summarised per age decade.

**Table 1:** PRESAGE test battery results for our group of age-adjusted normal listeners. Shown are the median and interquartile range [IQR] with sample size (n) for the combined dataset, alongside the linear regression intercept (β_0_), slope (β_1_), Spearman’s rank correlation coefficient (ρ), and corresponding p-values in relation to age and PTA. p-values were not adjusted for multiple comparisons. Significant p-values (p<0.05) are highlighted in bold.

| Variable | Median [IQR], $n$ | Age | | | | PTA | | | |
| --- | --- | --- | --- | --- | --- | --- | --- | --- | --- |
| | | $\beta_0$ | $\beta_1$ | $\rho$ | $p$-value | $\beta_0$ | $\beta_1$ | $\rho$ | $p$-value |
| ACALOS L15 (dB SPL) | 54.6 [50.9, 57.3], $n=55$ | 32.49 | 0.33 | 0.43 | **<0.01** | 52.36 | 0.17 | 0.12 | 0.399 |
| ACALOS L35 (dB SPL) | 92.0 [89.5, 94.0], $n=55$ | 83.58 | 0.11 | 0.16 | 0.254 | 91.93 | -0.11 | -0.14 | 0.294 |
| ACALOS DR (dB) | 36.7 [34.0, 40.3], $n=55$ | 51.08 | -0.21 | -0.35 | **<0.01** | 39.57 | -0.28 | -0.25 | 0.064 |
| ACALOS BLS (dB) | 7.5 [3.5, 10.9], $n=54$ | 7.65 | $2.71 \cdot 10^{-3}$ | 0.05 | 0.708 | 9.47 | -0.19 | -0.08 | 0.543 |
| TIN ERB<sub>500Hz</sub> (dB SNR) | 0.2 [-1.1, 0.7], $n=52$ | -3.74 | 0.05 | 0.28 | **<0.05** | -0.74 | 0.06 | 0.16 | 0.260 |
| TIN ERB<sub>2000Hz</sub> (dB SNR) | -2.5 [-3.9, -1.1], $n=52$ | -6.31 | 0.06 | 0.33 | **<0.05** | -2.96 | 0.04 | 0.16 | 0.265 |
| Matrix SRT SSN<sub>UOL</sub> (dB SNR) | -5.4 [-5.8, -5.0], $n=39$ | -9.20 | 0.06 | 0.65 | **<0.001** | -6.09 | 0.11 | 0.57 | **<0.001** |
| Matrix SRT SSN<sub>IDA</sub> (dB SNR) | -3.9 [-5.4, -2.1], $n=16$ | -10.64 | 0.11 | 0.39 | 0.132 | -5.28 | 0.16 | 0.39 | 0.139 |
| Matrix SRT<sub>norm</sub> SSN (dB SNR) | 1.7 [1.2, 2.8], $n=55$ | -2.74 | 0.07 | 0.58 | **<0.001** | 0.87 | 0.13 | 0.52 | **<0.001** |
| Matrix SRT ICRA5-250<sub>UOL</sub> (dB SNR) | -14.3 [-16.5, -13.4], $n=39$ | -26.89 | 0.19 | 0.49 | **<0.01** | -17.85 | 0.47 | 0.66 | **<0.001** |
| Matrix SRT ICRA5-250<sub>IDA</sub> (dB SNR) | -11.0 [-12.8, -8.3], $n=16$ | -22.52 | 0.19 | 0.42 | 0.102 | -14.34 | 0.36 | 0.60 | **<0.05** |
| Matrix SRT<sub>norm</sub> ICRA5-250 (dB SNR) | 5.1 [3.2, 6.5], $n=55$ | -7.28 | 0.19 | 0.49 | **<0.001** | 1.74 | 0.41 | 0.68 | **<0.001** |
| MEMR averaged threshold (dB SPL) | 86.7 [81.7, 90.0], $n=49$ | 88.04 | -0.02 | -0.04 | 0.804 | 87.48 | -0.09 | -0.08 | 0.595 |
| ECochG latency wave I (ms)<sup>1</sup> | 1.6 [1.5, 1.7], n=49 | 1.07 | 8.48·10<sup>-3</sup> | 0.52 | **<0.001** | 1.56 | 6.30·10<sup>-3</sup> | 0.19 | 0.189 |
| ECochG interpeak I-V (ms)<sup>1</sup> | 4.1 [4.0, 4.2], n=49 | 4.07 | 8.71·10<sup>-4</sup> | 0.03 | 0.838 | 4.09 | 4.76·10<sup>-3</sup> | 0.10 | 0.501 |
| ECochG interpeak III-V (ms)<sup>1</sup> | 1.9 [1.7, 1.9], n=53 | 1.84 | 1.20·10<sup>-4</sup> | -0.04 | 0.778 | 1.83 | 2.49·10<sup>-3</sup> | 0.03 | 0.808 |
| Caloric test SPV cold (°/s)<sup>2</sup> | 12.0 [9.0, 16.0], n=49 | 7.81 | 0.09 | 0.02 | 0.896 | 13.67 | -0.02 | 0.04 | 0.779 |
| Caloric test SPV warm (°/s) | 18.0 [10.8, 27.2], n=48 | 20.73 | -6.02·10<sup>-3</sup> | 0.03 | 0.855 | 22.74 | -0.27 | 0.01 | 0.968 |
| Caloric test UW (%)<sup>2</sup> | 4.2 [-7.7, 11.9], n=48 | -13.57 | 0.25 | 0.13 | 0.378 | -4.43 | 0.85 | 0.30 | **<0.05** |
| Caloric test DP (%) | -1.5 [-9.0, 13.8], n=48 | 34.11 | -0.53 | -0.19 | 0.193 | -0.47 | 0.04 | 0.05 | 0.737 |
| vHIT gain Anterior | 1.0 [0.9, 1.0], n=54 | 1.16 | -3.15·10<sup>-3</sup> | -0.21 | 0.120 | 0.99 | -4.33·10<sup>-3</sup> | -0.18 | 0.181 |
| vHIT gain Lateral | 1.0 [0.9, 1.0], n=54 | 0.99 | -5.17·10<sup>-5</sup> | 0.08 | 0.563 | 1.01 | -3.69·10<sup>-3</sup> | -0.14 | 0.325 |
| vHIT gain Posterior | 0.9 [0.8, 0.9], n=53 | 0.99 | -2.46·10<sup>-3</sup> | -0.21 | 0.137 | 0.87 | -4.91·10<sup>-3</sup> | -0.07 | 0.602 |
| vHIT AR Anterior (%) | -0.5 [-3.4, 2.6], n=54 | 2.51 | -0.04 | 0.12 | 0.389 | -0.90 | 0.12 | 0.30 | **<0.05** |
| vHIT AR Lateral (%) | -0.5 [-3.4, 3.4], n=54 | 6.17 | -0.10 | -0.12 | 0.398 | 1.10 | -0.13 | -0.19 | 0.177 |
| vHIT AR Posterior (%) | -1.1 [-3.5, 0.8], n=52 | -4.16 | 0.05 | -0.09 | 0.538 | -0.95 | -3.64·10<sup>-3</sup> | -0.09 | 0.536 |
| cVEMP EMG-normalised P-N amplitude (µV) | 0.7 [0.5, 0.9], n=52 | 0.90 | -1.64·10<sup>-3</sup> | -0.01 | 0.934 | 0.79 | 7.30·10<sup>-4</sup> | 0.01 | 0.941 |
| cVEMP AR (%) | 0.0 [-10.8, 11.4], n=50 | -18.41 | 0.29 | 0.22 | 0.131 | -1.51 | 0.19 | -0.03 | 0.823 |
| oVEMP EMG-normalised P-N amplitude (µV) | 0.6 [0.3, 0.8], n=48 | 1.03 | -5.84·10<sup>-3</sup> | -0.18 | 0.220 | 0.64 | 1.54·10<sup>-3</sup> | -0.06 | 0.698 |
| oVEMP AR (%) | 4.9 [-3.6, 24.1], n=46 | 14.18 | -0.13 | -0.10 | 0.500 | 12.75 | -0.92 | -0.23 | 0.122 |

<sup>1</sup>only latencies measured at 115 dB SPL were included; <sup>2</sup>monothermal SPVs were excluded.

### Audiological functional measures

#### Adaptive categorical loudness scaling

Figure 2 shows scatterplots of ACALOS outcome variables as a function of age and PTA. Data points are colour-coded by age to aid visual interpretation, with UOL participants indicated by a black outline. The assessed variables are the levels L15 (soft category) and L35 (loud category; both in first column), dynamic range (DR, second column), and binaural loudness summation (BLS, third column). For binaural loudness summation, the results obtained at L15 and L35 were averaged before analysis since no significant difference between the two categories was found (β = 0.95, SE = 0.87, 95% CI [-0.79, 2.69], t(53) = 1.10, p = 0.28), as expected for IFNoise according to van Beurden et al. (2018). Spearman’s rank correlations confirmed a significant positive association between age and L15 (ρ = 0.43, p < 0.001), but not with L35 (ρ = 0.16, p = 0.25). Neither L15 nor L35 were significantly correlated with PTA (both p’s > 0.05). The age-related increase for L15, in the absence of a correlation with PTA, suggests that loudness perception in normal-hearing adults may be influenced by age independently of audiometric sensitivity. DR showed a significant negative correlation with age (ρ = -0.35, p < 0.05), and a weaker, non-significant trend with PTA (ρ = -0.25, p = 0.064). No significant correlations were found between BLS and either predictor (both p’s > 0.05), though substantial inter-individual variability was observed. This translated to an estimated L15 increase of ∼3 dB per age decade, whereas L35 remained largely stable with a median of 92 dB SPL (IQR [89.5, 94.0] dB SPL). DR declined by ∼2 dB per decade, while BLS remained stable at a median of 7.5 dB (IQR [3.5, 10.9] dB), with positive BLS values indicating a higher loudness sensitivity to binaural than to monaural broadband stimuli, but generally lower than found by van Beurden et al. (2018).

**Figure 2.**
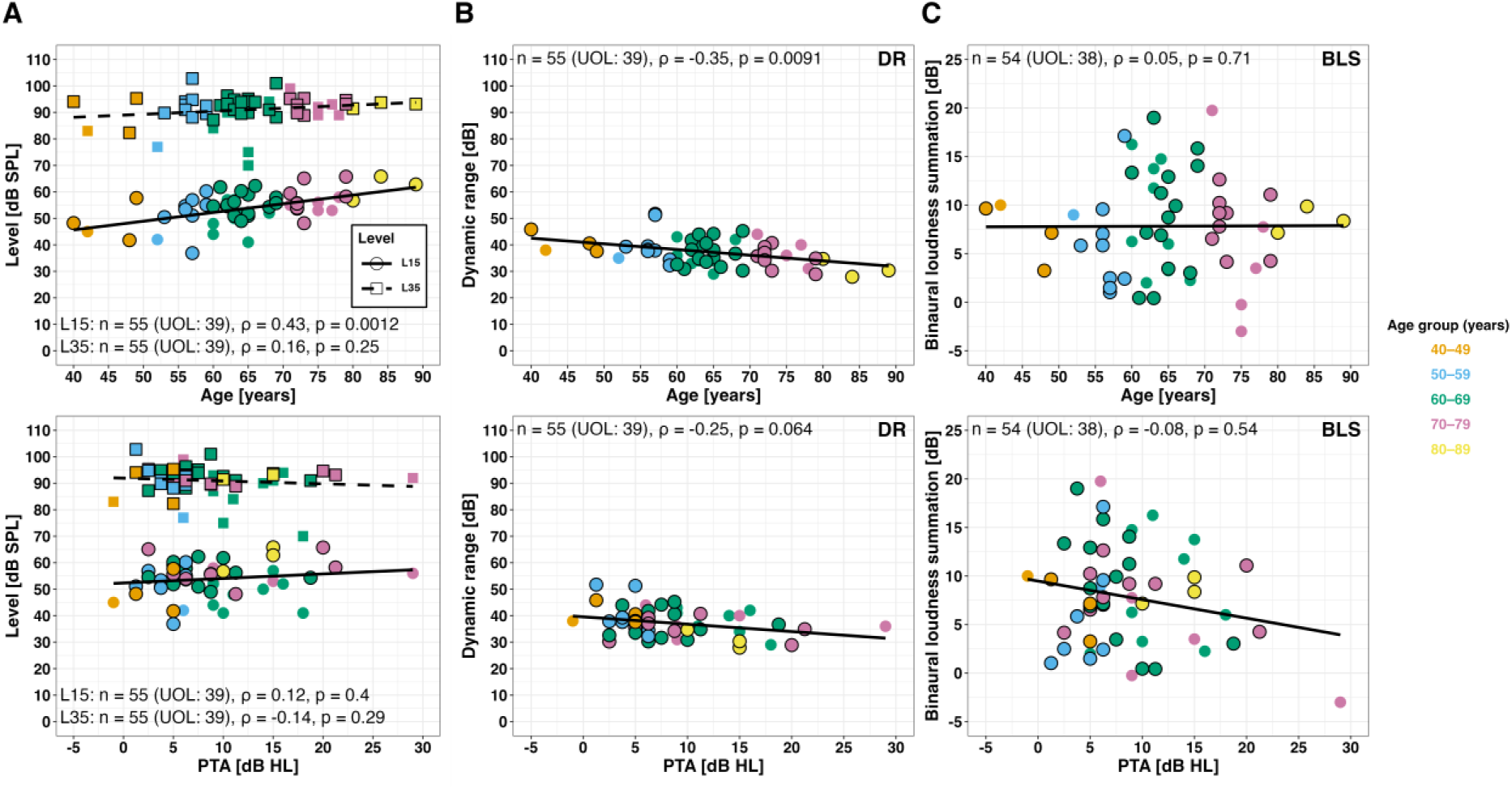
Adaptive Categorical Loudness Scaling (ACALOS) results as a function of age (top) and PTA (bottom). Panel A shows estimated levels at L15 (circles, solid lines) and L35 (squares, dashed lines); in panels B and C, symbols/lines represent dynamic range (DR) and binaural loudness summation (BLS), respectively. UOL participants are shown with black-outlined symbols and IDA participants without an outline; colours indicate age groups. Correlations and regression lines were calculated on the combined dataset across centres. Spearman’s rank-order correlation coefficients (ρ), corresponding p-values, and sample sizes (n) are shown within each panel. The same plotting conventions apply to Figures 3–11, except for Figure 6.

#### Tone-in-noise

Figure 3 shows TIN detection thresholds over age and PTA, assessed at 500 Hz and 2 kHz. Spearman’s rank correlations confirmed a significant positive association with age at both frequencies (500 Hz: ρ = 0.28, 2 kHz: ρ = 0.33, both p < 0.05), whereas correlations with PTA were weaker and not significant (both ρ = 0.16, p > 0.26). This translated to an estimated increment in detection thresholds of about 1 dB per age decade at both frequencies. Median thresholds were close to 0 dB SNR at 500 Hz (0.2 dB SNR; IQR [-1.1, 0.7] dB SNR) and slightly lower at 2 kHz (−2.5 dB SNR; IQR [-3.9, -1.1] dB SNR). Noticeable inter-individual variability was observed, particularly at 2 kHz, where participants of comparable age or PTA still showed a wide spread in thresholds, for example from -8 to 0 dB SNR in the 60-69-year age group.

**Figure 3.**
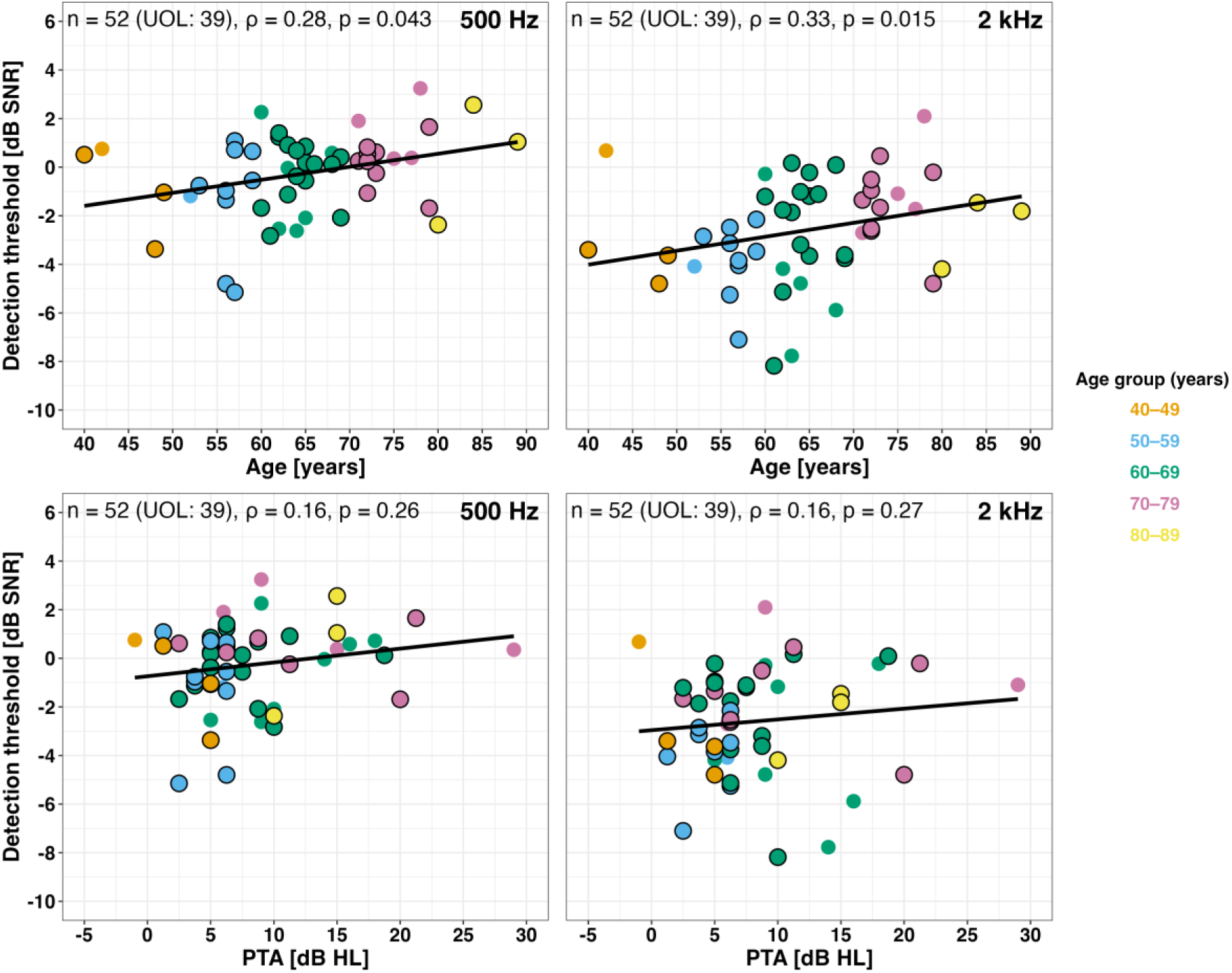
Tone-in-noise (TIN) detection thresholds at 500 Hz (left) and 2 kHz (right) as a function of age (top) and better-ear PTA (bottom).

#### Matrix sentence test

Reference-normalised SRTs (SRT_norm_) for the Matrix sentence test, pooled across centres, are shown in Figure 4 as a function of age and PTA. SRT_norm_ values increased significantly with both age and PTA in both noise types. For SSN, SRT_norm_ values increased by approximately 1 dB per 10 years (ρ = 0.58, p < 0.001) and per 10 dB HL (ρ = 0.52, p < 0.001). In ICRA5-250, the slope was steeper, with SRT_norm_ values increasing by approximately 2 dB per 10 years (ρ = 0.49, p < 0.001) and 4 dB per 10 dB HL (ρ = 0.68, p < 0.001). Notably, listeners with comparable PTA performed similarly regardless of age, indicating that the observed age effects are primarily mediated by hearing sensitivity rather than biological age. The language-specific SRTs were close to the respective published reference values for both languages and noise conditions (Wagener et al., 1999a, b, c; Hochmuth et al., 2015; Jansen et al., 2012) for the youngest participants at about 40 years, as represented by SRT_norm_ values close to 0 dB in Figure 4. Performance was less variable in SSN, with an IQR of 1.6 dB on the pooled SRT_norm_ data. In contrast, SRTs in fluctuating noise (ICRA5-250) showed broader inter-individual variability (IQR of 3.3 dB).

**Figure 4.**
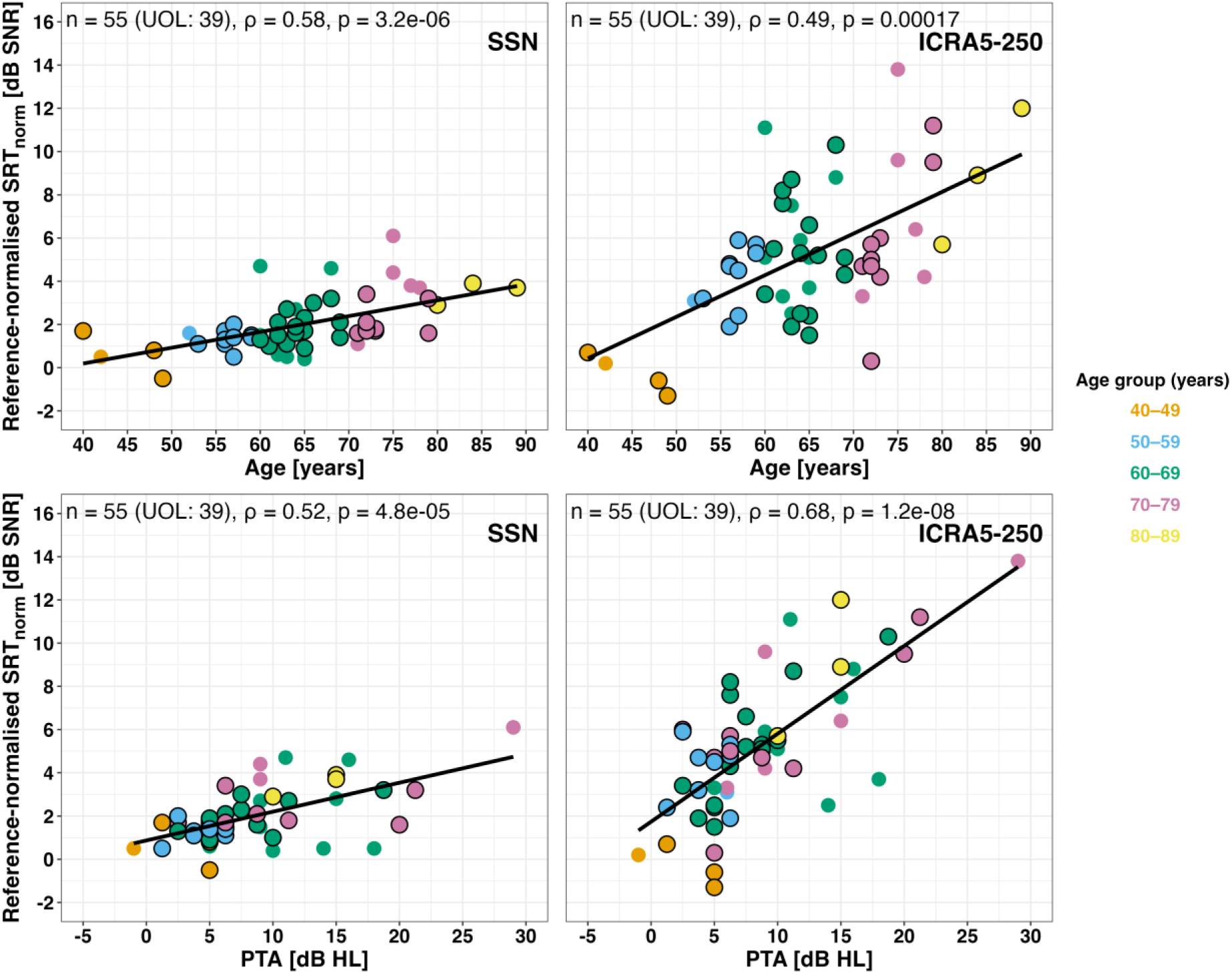
Reference-normalised speech recognition thresholds (SRTnorm) from the Matrix sentence test, pooled across centres, as a function of age (top) and better-ear PTA (bottom). Results are shown separately for test-specific stationary noise (SSN; left) and fluctuating noise (ICRA5-250; right).

### Audiological physiological measures

#### Middle-ear muscle reflex

Figure 5 shows middle-ear muscle reflex (MEMR) thresholds, defined as the mean level across 500 Hz, 1 kHz, and 2 kHz. A MEMR was classified as present if at least one of the three tested frequencies (500 Hz, 1 kHz, 2 kHz) elicited a response. This criterion was met in most participants (49/55, 89.1%), while 6 participants (10.9%) showed no triggered (NT) MEMR (grey points). Spearman’s rank correlations showed no significant associations with either age or PTA (p > 0.05), indicating that MEMR thresholds were relatively stable across age and degree of hearing loss in participants with age-normal PTA, with a median of 86.7 dB SPL (IQR [81.7, 90.0] dB SPL). MEMR averaged thresholds showed large inter-individual variability, with some very high values despite age-normal PTA.

**Figure 5.**
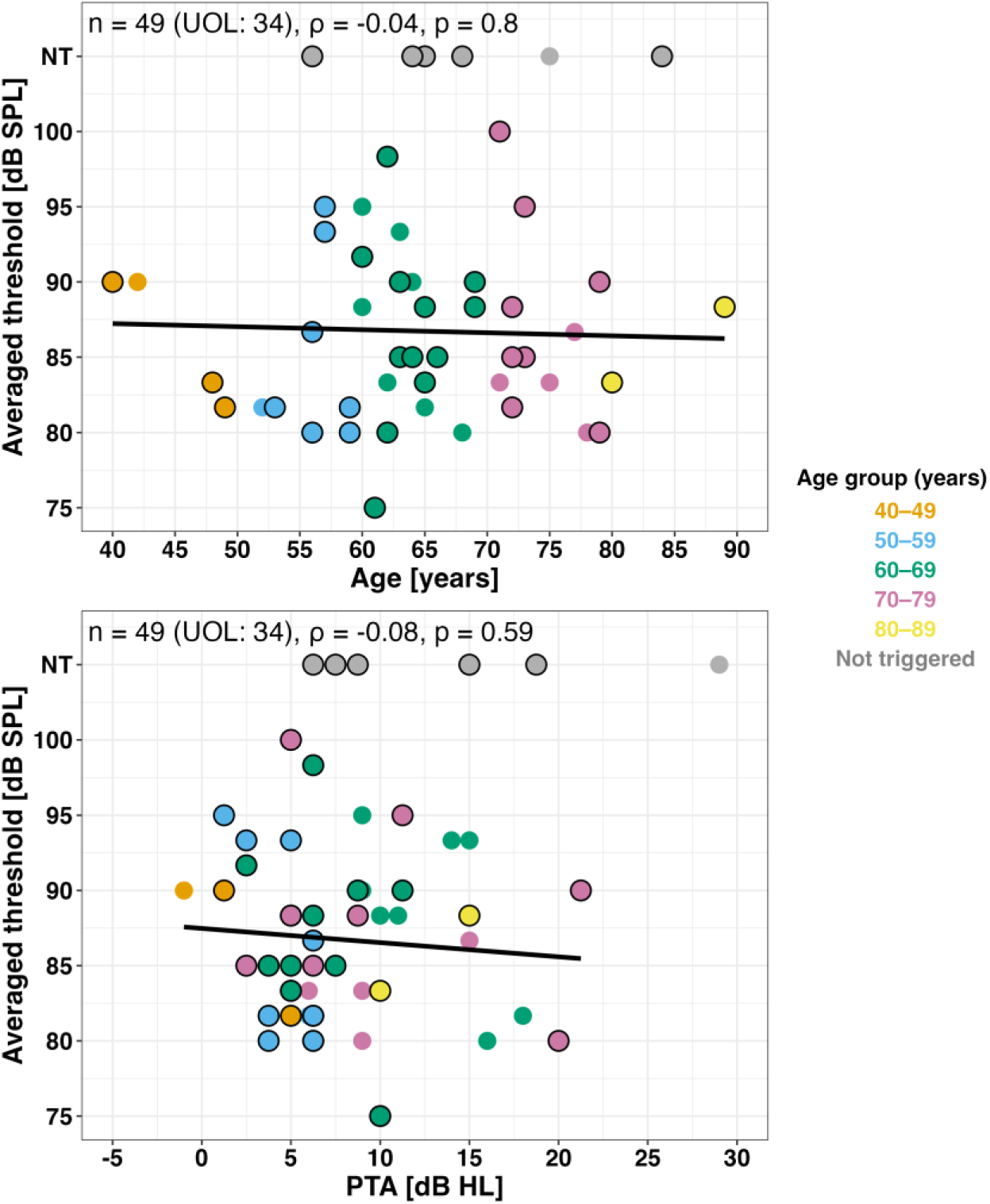
MEMR thresholds averaged across 500 Hz, 1 kHz, and 2 kHz as a function of age (top) and better-ear PTA (bottom). Not triggered points (NT) are shown in grey.

#### Otoacoustic emissions

Figure 6 displays frequency-specific and global pass proportions of present otoacoustic emission responses. Panel A presents TEOAE responses at 1–5 kHz, and panel B presents DPOAE responses at 1, 2, and 4 kHz. The pass criterion was defined as having at least three present responses out of the five measured frequencies for TEOAEs and at least two of the three measured frequencies for DPOAEs. For TEOAEs, this criterion was fulfilled by 45 of 53 participants (85%). The participants with absent TEOAE were all older than 57 years. In two additional participants, TEOAEs responses could not be obtained. The proportion of present responses was highest at 2 and 3 kHz (87% in each) intermediate at 1 kHz (68%) and 4 kHz (51%), and lowest at 5 kHz (23%). This pattern of weaker responses with increasing frequencies is characteristic of click-evoked TEOAEs in normal-hearing adults (Kemp, 2002; Hoth et al., 2010).

**Figure 6.**
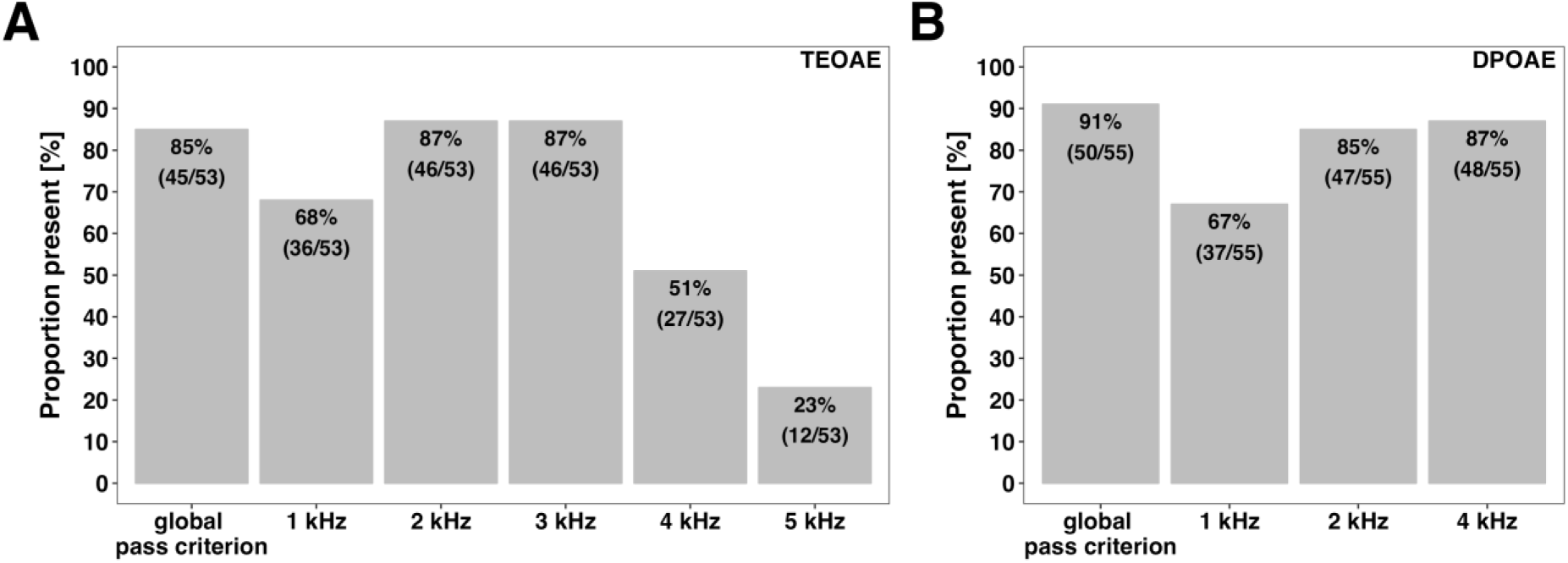
Proportion of global and frequency-specific presence of TEOAE (A) and DPOAE (B).

As for DPOAEs, 41 participants (75.5%) were measured only at the lower level (L1 = 65 dB SPL, L2 = 60 dB SPL), and 14 participants (25.5%) were additionally measured at the higher level (both L1 and L2 at 68 dB SPL). The pass criterion was fulfilled by 50 of 55 participants (91%). One of the five participants without globally present DPOAEs had been kept at 65 dB SPL because present responses were detected at 4 kHz and at additional IDA-specific frequencies (1.5, 3, and 5 kHz), which fulfilled the centre-specific pass rule, despite only one present response within the shared frequency range across centres (1, 2, and 4 kHz). Interestingly, the participants with globally absent DPOAEs were all older than 60 years. Three out of five participants showed consistent absence of both TEOAE and DPOAE. The frequency-specific proportions of present responses were highest at 4 and 2 kHz (87% and 85%, respectively) and lowest at 1 kHz (67%). The reduced presence at f2 = 1 kHz is likely due to the limited ability of the apical outer hair cells (OHC) to synchronise in phase. Hence, the analysed frequency of 2*f1-f2 = 656 Hz represents a frequency close to the lower limit of DPOAE measurements.

#### Electrocochleography

Figure 7 shows electrocochleography (ECochG) latencies for wave I, and interpeak intervals I-V and III-V. Wave I was absent in four participants (grey circles), consistent with literature describing it as the first component to diminish with age (Qian et al., 2021; Oku & Hasegewa, 1997). Analyses were carried out on data measured at 115 dB SPL (90 dB nHL), with data from 49 participants for wave I and interpeak I-V, and from 53 participants for interpeak III-V. Spearman’s rank correlations revealed a significant positive association of wave I latency with age (ρ = 0.52, p < 0.001), corresponding to an increase of about 0.1 ms per decade. No significant association with PTA was observed for wave I (p = 0.19). For the interpeak intervals I-V and III-V, no significant correlations were found with either age or PTA (all p’s > 0.5). Median interpeak intervals were 4.1 ms (IQR [4.0,4.2] ms) for I-V and 1.9 ms (IQR [1.7,1.9] ms) for III-V. Hence, brainstem generators and circuits are remarkably age-insensitive in the control group, as interpeak latency I-V was missing only when wave I was missing. Although there is limited literature reporting ECochG interpeak interval I–V and III–V in older, normal-hearing adults, normative ABR data provide a useful, though not perfect, comparison. For example, Minaya & Atcherson (2015) found that ECochG action potential latency closely matches ABR wave I latency, suggesting that ABR interpeak intervals might approximate ECochG conduction times. Normative ABR I–V latency values in adult populations often cluster around ∼4.0–4.2 ms, consistent with our median value of 4.1 ms (e.g., Sanfins et al., 2022).

**Figure 7.**
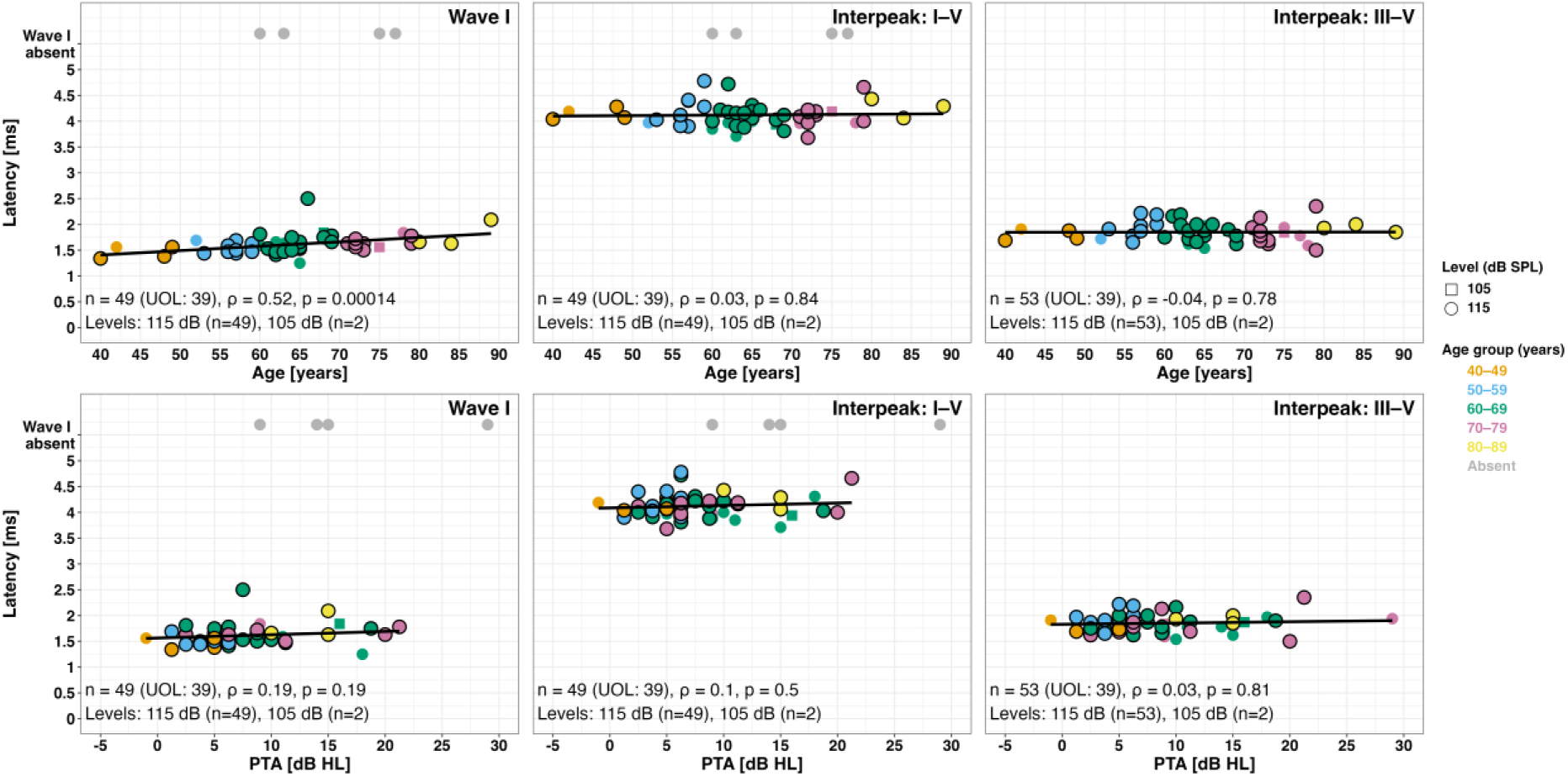
ECochG latencies for wave I, interpeak I-V, and III-V as a function of age (top) and better-ear PTA (bottom). Grey circles indicate absent wave I. Regression lines and correlations were calculated only for recordings at 115 dB SPL.

### Vestibular physiological measures

#### Video head impulse test

Video head impulse test (vHIT) gains for the anterior, lateral, and posterior semicircular canals are shown in Figure 8A. Four data points were excluded from the analysis as they could not be reliably measured (grey circles). One participant yielded not-measurable (NM) responses for all three canals due to movement artefacts, while in another, the posterior canal was NM due to poor pupil tracking. No significant associations with either age or PTA were found (all p’s > 0.18), with median gains of 1.0 (IQR [0.9,1.0]) for anterior and lateral canals, and 0.9 (IQR [0.8,0.9]) for the posterior canal. Considering the cut-off of 0.8 for lateral and 0.7 for anterior and posterior canals (Strupp et al., 2019, Goto et al., 2024), several participants showed lower than normal vHIT gain, especially for the posterior canal.

**Figure 8.**
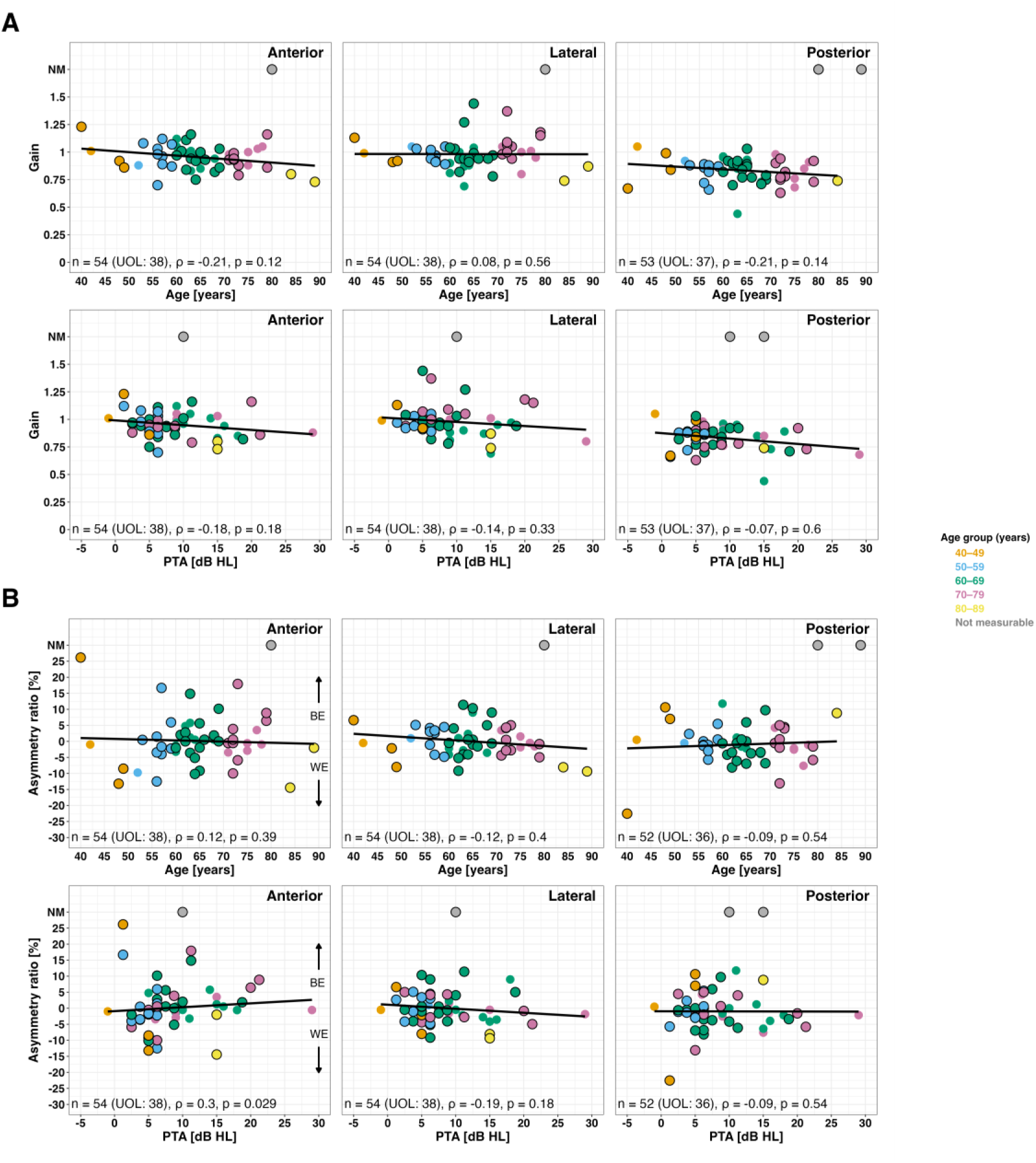
vHIT gains (panel A) and gain asymmetry ratios (panel B) for anterior, lateral, and posterior semicircular canals as a function of age (top) and better-ear PTA (bottom). (A) Grey circles denote not measurable (NM) data.

Figure 8B shows vHIT gain asymmetry (AR) for the anterior, lateral, and posterior canals. The asymmetry ratio (AR) sign is defined relative to the PTA better ear, with positive values indicating a stronger response toward the PTA better-ear side. No significant associations between AR and age were found (all p’s ≥ 0.39), with ARs centred near zero and medians of -0.5% (IQR [-3.4,2.6] % for anterior; IQR [-3.4,3.4] % for lateral) and -1.1% (IQR [-3.5,0.8] %) for the posterior canal. For PTA, a small but significant positive association was observed for the anterior canal (ρ = 0.30, p < 0.05), corresponding to about 1% increase in AR per 10 dB HL, while lateral and posterior canals showed no significant associations (p = 0.18 and p = 0.54, respectively).

#### Vestibular-evoked myogenic potentials

Figures 9A and 10A show EMG-normalised peak-to-peak (P1-N1) amplitudes for cervical and ocular VEMPs, respectively, and Figures 9B and 10B the corresponding asymmetry ratios (ARs), computed for participants with complete bilateral responses, as a function of age and PTA.

**Figure 9.**
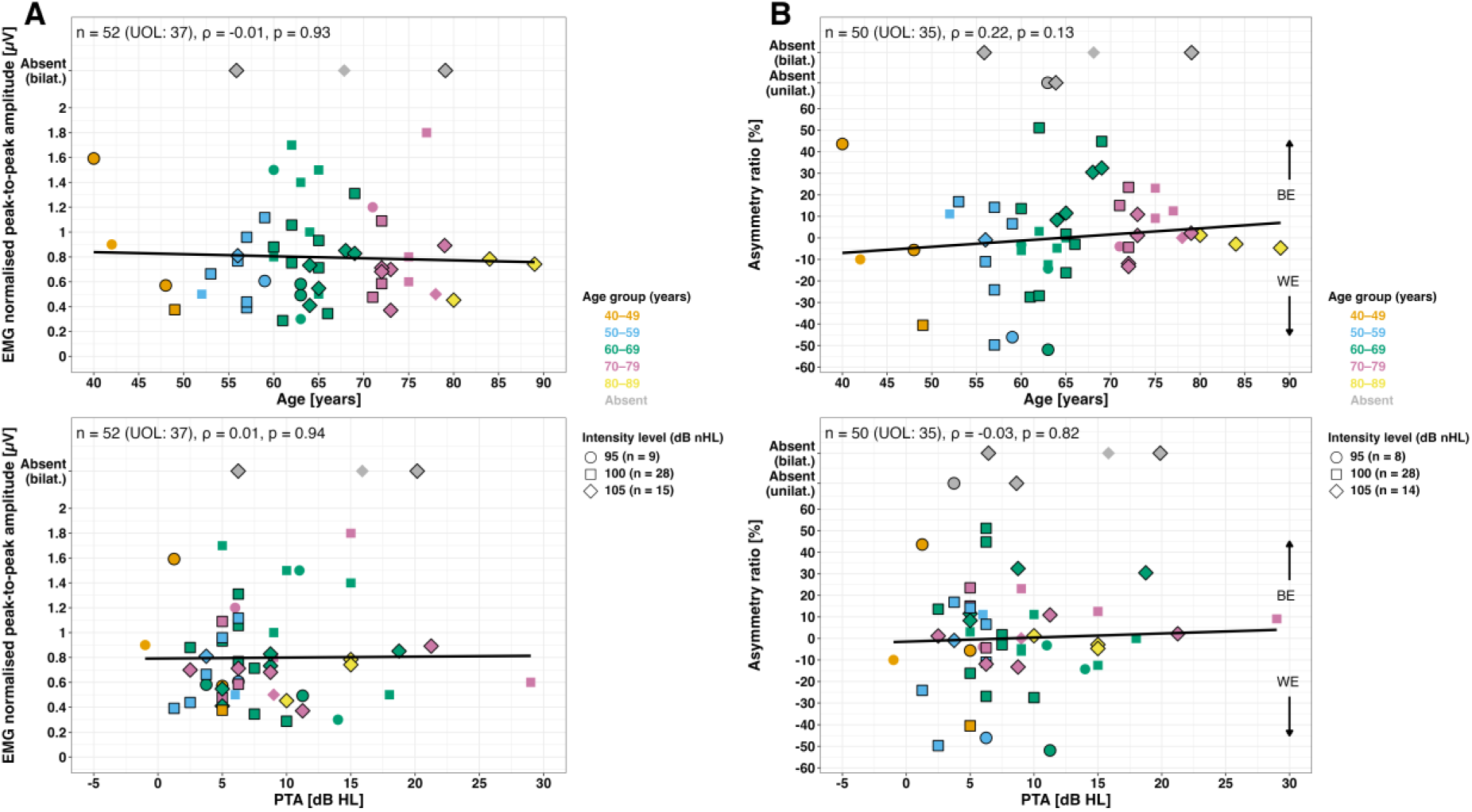
EMG-normalised cVEMP peak-to-peak amplitudes (panel A) and asymmetry ratios (panel B) as a function of age (top) and better-ear PTA (bottom). Symbols indicate the stimulus intensity (dB nHL) at which a measurable response was obtained (about 5 dB above threshold). For statistical analysis, responses for all levels were combined. Grey symbols denote bilateral or unilateral absent responses.

**Figure 10.**
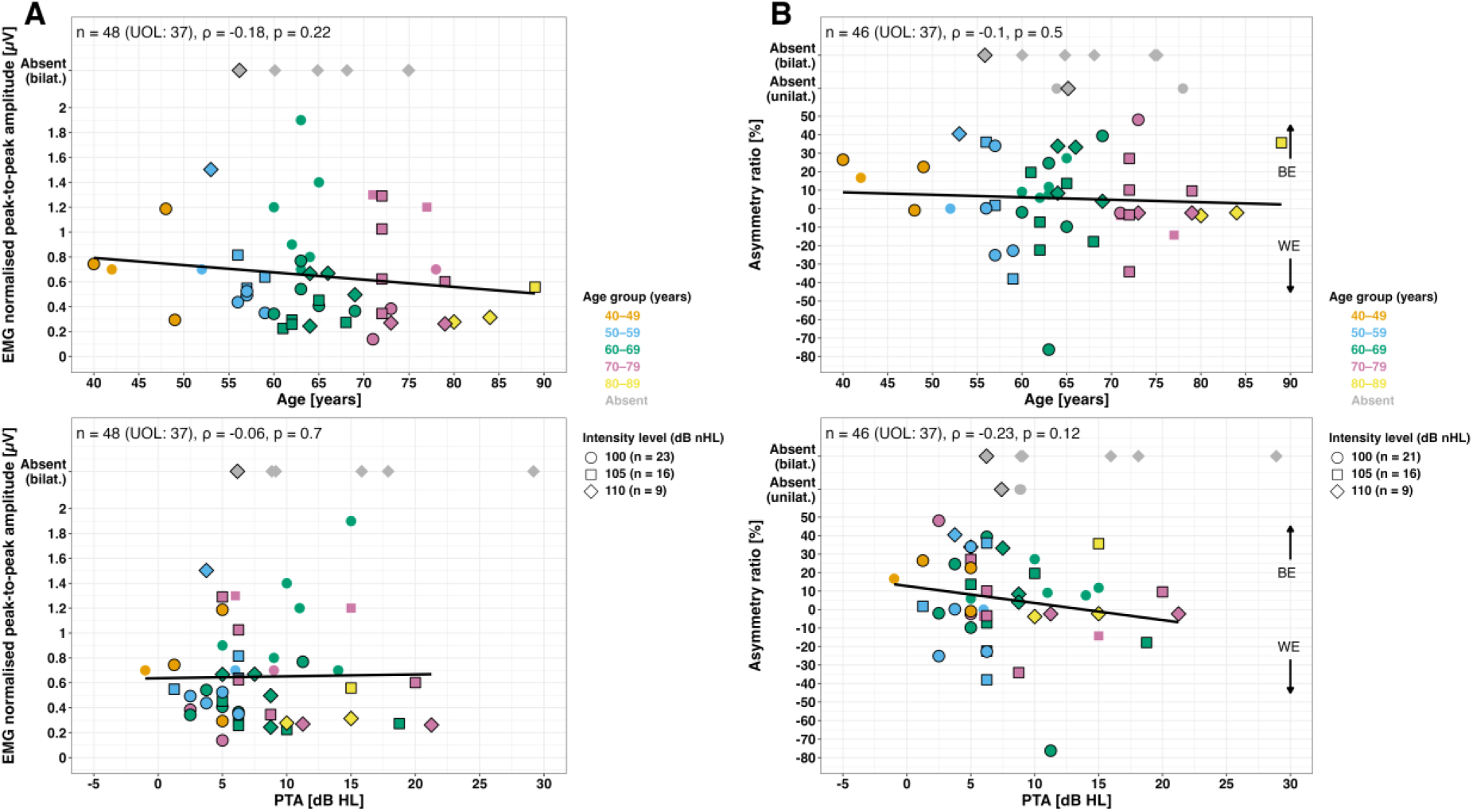
EMG-normalised oVEMP peak-to-peak amplitudes (panel A) and asymmetry ratios (panel B) as a function of age (top) and better-ear PTA (bottom). Symbols indicate the stimulus intensity (dB nHL) at which a measurable response was obtained (about 5 dB above threshold). Regression lines and correlations were computed for all responses irrespective of stimulus intensity. Grey symbols denote bilateral or unilateral absent responses.

For cVEMPs, bilateral responses were absent in three participants and unilateral responses in two (both in the PTA worse ear). For the remaining 52 participants, most responses were obtained at 100 ± 5dB nHL, with 17% at 95 dB, 54% at 100 dB, and 29% at 105 dB nHL. No significant correlations were found between P1-N1 amplitudes and either age or PTA (both p’s > 0.9), with a median normalised amplitude of 0.7 µV (IQR [0.5,0.9] µV). Likewise, no significant associations were found between cVEMP asymmetry ratios for either age or PTA (both p’s > 0.1) with median AR of 0.0% (IQR [-10.8, 11.4] %). For oVEMPs, bilateral responses were absent in six participants and unilateral responses in three (two in the PTA worse ear and one in the better ear). Of the remaining 48 participants, approximately half of the responses (48%) were obtained at 100 dB, one third (33%) at 105 dB and 19% at 110 dB. As with cVEMPs, no significant associations were found between oVEMP peak-to-peak amplitudes or ARs and either age or PTA (both p’s > 0.7 and > 0.12, respectively), with a median peak-to-peak amplitude of 0.6 µV (IQR [0.3, 0.9] µV) and a median AR of 4.9% (IQR [-3.6, 24.1] %). Hence, all participants can be considered normal based on the normalised amplitude in their PTA better ear. For example, Pandey et al. (2025) reported a median normalised cVEMP amplitude of about 0.4 µV (IQR 0.3 µV) for participants with normal hearing and vestibular function. Asymmetry ratios below approximately 20% are considered normal (Nguyen et al., 2010; Isaradisaikul et al., 2012). Hence, several participants exhibited asymmetric vestibular results in VEMPs, both in the direction of the better or worse PTA ear.

#### Caloric test

Figure 11A shows caloric slow-phase velocity responses (SPVs) for cold and warm irrigations, and Figure 11B shows the corresponding asymmetry metrics unilateral weakness (UW) and directional preponderance (DP) as a function of age and PTA. For one participant, only cold irrigation recordings were obtained because of intolerance (marked ‘M’ for monothermal). For completeness, for this participant, UW was estimated using the monothermal caloric asymmetry (MCA) formula but excluded from the analyses, as cold MCA has limited diagnostic reliability (Lightfoot et al., 2009).

**Figure 11.**
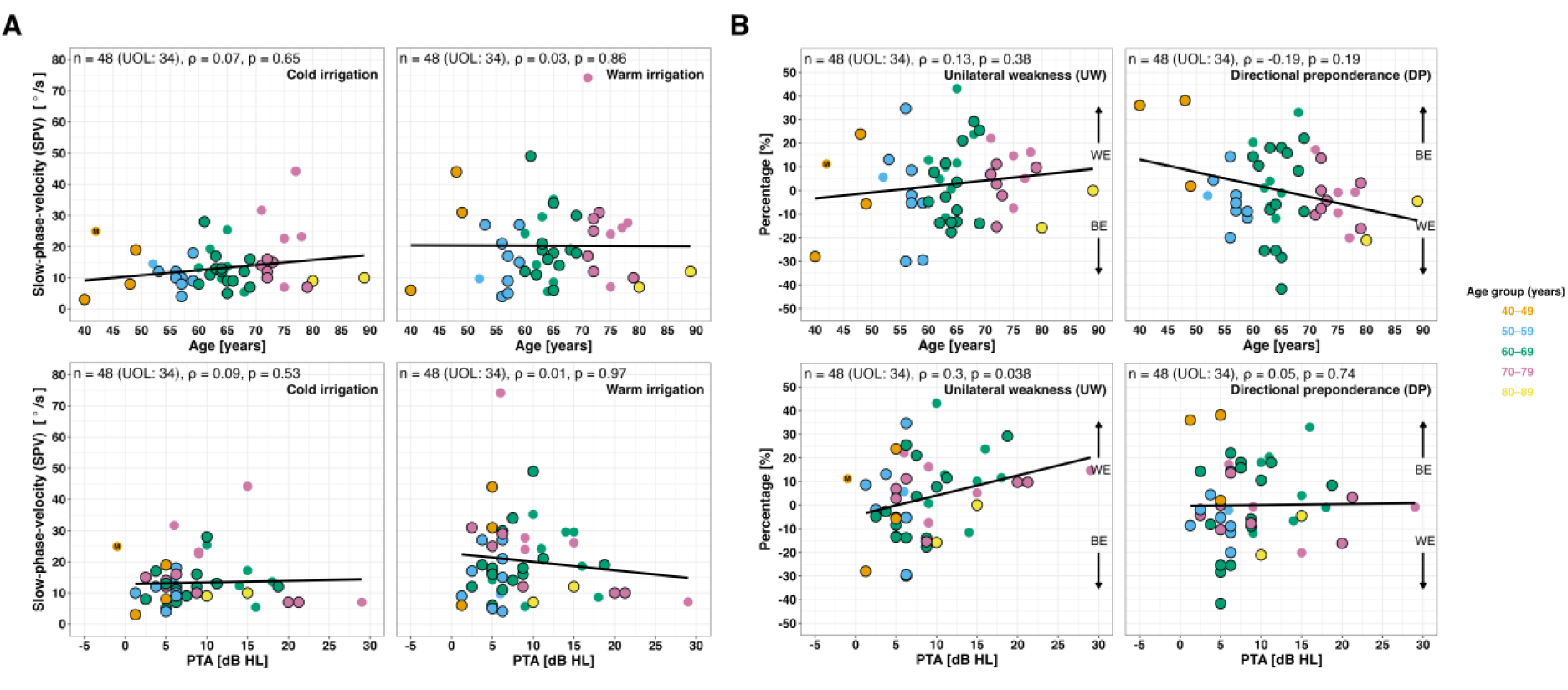
Slow phase velocity (SPV) for cold and warm irrigation (panel A) and corresponding asymmetry metrics of unilateral weakness (UW) and directional preponderance (DP) in panel B as a function of age (top) and better-ear PTA (bottom). One monothermal data point was excluded from the correlation and regression analyses (marked with ‘M’).

SPVs showed no significant associations with either age or PTA (p > 0.53), with median velocities of 12.0 °/s (IQR [9.0,16.0] °/s) for cold and 18.0 °/s (IQR [10.8, 27.2] °/s) for warm irrigation. Similarly, no significant correlations with age were found for either UW or DP (p > 0.19). As for PTA, UW showed a small positive association (ρ = 0.30, p < 0.038), corresponding to an ∼8.5% increase in UW per 10 dB HL increase in the better ear PTA. Since positive UW reflects a relative weakness of the PTA worse ear, this pattern indicates a tendency toward smaller SPVs on the side with poorer hearing. In contrast, DP showed no significant correlation with PTA (ρ = 0.05, p = 0.74). Median UW and DP were 4.2% (IQR [-7.7,11.9] %) and -1.5% (IQR [-9.0,13.8] %), respectively. Strupp et al. (2020) reported reference values used across different labs. Mean peak slow phase velocity of caloric-induced nystagmus was 18.65°/s (12–30°/s) for warm irrigation and 18.21°/s (10–25°/s) for cold irrigation. All participants showed caloric asymmetry values below the pathological side difference reported by Strupp et al. (2020), whose mean was 25.93% (range: 17.7%–40%).

## Discussion

This study introduced a cross-domain test battery for comprehensive characterisation of hearing loss based on functional, physiological, and vestibular tests (objective 1). The comparability of data collected in Germany and France was evaluated across all tests, and univariate relationships between measured outcomes and participants’ age as well as PTA were examined. A reference group of participants from both countries, characterised by age-normal hearing thresholds according to ISO 7029 (2017), was included. This group was selected to establish and explore fundamental relationships that will be essential for interpreting data from hearing-impaired participants in future studies. Focusing on this (nearly) normal-hearing group allows controlled investigation of inter-centre data comparability and provides insights into various hearing-related domains beyond audibility. Importantly, the test battery is designed to capture complementary aspects of hearing function, with the potential to reveal subclinical or atypical results even in participants with normal-hearing PTA.

### Data comparability across centres

The second objective of this study was to assess the comparability of data across the two test centres (UOL and IDA), as an important prerequisite for combined interpretation of the results. In the audiological-functional domain, centre effects were identified for the loudness scaling level variables L15 and L35. No centre effect was identified in the audiological-physiological domain. For the vestibular-physiological tests, centre effects occurred for slow-phase velocity (SPV) at cold stimulation in the caloric test, for the lateral gain in vHIT, and for the normalised cVEMP and oVEMP amplitudes. All data were analysed in combination after careful assessment of the underlying reasons for the centre differences, making pooling plausible. The justification for combining data included: i) known differences between measurement equipment used at UOL and IDA (particularly for vestibular tests), ii) centre effects being smaller than inter-individual variability reported in the literature (e.g., ACALOS), iii) centre effects affecting only single variables within a test.

For the Matrix sentence test, the centre effect observed in the language-specific SRTs was expected, as the German and French test versions differ systematically in their normative SRTs by about 1 dB for stationary noise (Wagener et al., 1999a, b, c; Jansen et al., 2012) and about 3 dB in fluctuating noise (Hochmuth et al., 2015; Jansen et al., 2012). Normalisation with published language- and noise-dependent normative values (SRT_norm_) was appropriate because the psychometric function slopes in stationary noise are similar (17.1 %/dB for the German, and 14.0 %/dB for the French version; Kollmeier et al., 2015), and both tests targeted a 50% speech recognition score. Additional considerations for combining Matrix test results would be required to compare data across Matrix tests with large differences between slopes or different target intelligibility. After normalisation, no centre effect was found in either noise type, indicating that the two language-specific versions yield comparable outcomes once their test-specific reference differences were accounted for. Consequently, SRT_norm_ values from both centres were pooled for further analyses.

Data for the normalised VEMP amplitudes were combined for analysis despite the identified strong centre effect (especially for oVEMP with p = 0.001 and Cohen’s d = 1.699). The datasets collected with different measurement devices (Eclipse and Neurosoft) spanned a similar value range, consistent with the expectation that these devices yield comparable results despite differences in EMG averaging (van Tilburg et al., 2019). The large overall value range (0.2 to 1.8) demonstrates a high variability, comparable to the variability of cVEMP normalised amplitudes as reported by Agrawal et al. (2013). The variability would have been expected to be even larger without normalisation with the EMG amplitude (van Tilburg et al., 2014). Given the small sample size of the matched groups for centre effect analysis (n=16 per centre), and considering that matching was based on PTA and age, while the tests in the test battery assess complementary auditory and vestibular domains, it is likely that the high variability in normalised VEMP amplitudes resulted in an apparent centre effect despite assumed comparability.

ACALOS data were analysed in combination since the observed centre effect of 5 dB fell within the intra-subject standard deviation for normal-hearing participants, which was reported by Oetting et al. (2014) as about 10 dB for lower and 4 dB for higher loudness categories. Thus, the estimated centre effect and the test-retest reliability cannot be disentangled in the data. Additionally, the same software and measurement devices (including calibration) were used at both centres, and the procedure is not language-dependent. Hence, factors beyond individual variability are not assumed to have influenced the results. These tests provide examples for our reasoning for combining data for analysis, which extends to all other variables showing a centre effect. For most tests, centre effects were smaller and visually easier to interpret, often affecting only single variables. In summary, the findings indicate that the proposed cross-domain test battery is feasible to be employed in multicentre studies. Nevertheless, careful harmonisation of measurement conditions and parameters, as well as a thorough understanding of the properties and comparability of the employed equipment, are essential.

### Audibility effects

The third objective of this study was to assess audibility effects with respect to the better-ear PTA. The inclusion criteria limited the PTA range to 40 dB HL for the oldest age, while most participants exhibited a PTA below 20 dB HL. Hence, the purpose was to investigate associations within PTAs better than the age-dependent median. In the audiological-functional domain, an audibility effect was identified for the Matrix test, whilst in audiological-physiological tests, no PTA dependencies were observed. In the vestibular domain, audibility effects emerged only for the asymmetry measures of the caloric test (UW) and the vHIT, specifically for the anterior canal asymmetry.

For the Matrix test, highly significant PTA effects (p<0.001) were found in the reference-normalised SRTs pooled across centres for both stationary and fluctuating noise. The observed PTA dependency in stationary noise replicates previous findings for the German matrix test by Wardenga et al. (2015). The larger inter-individual variability in fluctuating compared to stationary noise is in line with Wagener & Brand (2005). In future work, speech recognition modelling could help explain which parts of the variability relate to audibility and which to possible supra-threshold deficits, as reflected, e.g., in tone-in-noise detection thresholds (Schädler et al., 2020; Hülsmeier et al., 2022).

The remaining PTA effects were weaker, mostly reaching a significance level of 0.05. The PTA effect observed for the UW in the caloric test was unexpected from literature (Schubert et al., 2022). Given the symmetric PTA between ears, both vestibular systems should be similarly affected by higher PTA if a correlation exists, and hence, not cause increasing asymmetry. Similarly, the PTA effect on the vHIT anterior canal asymmetry was unexpected, but may have been driven by a small number of datapoints at higher PTAs.

Despite normal-hearing PTAs, several tests showed abnormal or atypical results in a substantial proportion of the participants, for example, tone-in-noise detection, binaural loudness summation, or middle-ear muscle reflex. It should be noted that robust evaluation of PTA effects requires data covering a broader PTA range. However, the primary aim of the present study was to identify effects that should be considered when interpreting results for participants classified as normal-hearing based on their PTA.

### Age effects

Significant age effects, also corresponding to the third objective of this study, were identified for all audiological-functional tests (ACALOS, Matrix test, TIN), while in the audiological-physiological domain, age effects were observed only for the ECochG wave I latency. No significant age effect was identified for any of the vestibular tests.

For ACALOS, the level at which sounds are perceived as soft (L15) increased with age (about 3 dB per age decade), accompanied by a reduction in dynamic range. The L15 effect has not yet been reported in literature, while a PTA dependency rather than age would have been expected due to the lower part of the loudness curve being influenced by the hearing threshold (Oetting et al., 2014). The observed dynamic range effect directly depends on the L15 effect, as it is calculated as the difference between L35 and L15. Further analyses across a broader PTA range are required to disentangle potential interactions between age and PTA. For this, leveraging large-scale public datasets such as the Oldenburg Hearing Health Record (OHHR), which includes ACALOS measurements in a cohort of 581 participants (aged 18–86 years), could enable a more precise characterisation of age-dependent loudness growth and improve the robustness of our findings (Jafri et al., 2025).

For the Matrix test, both noise conditions exhibited a significant age effect. For the German Matrix test in stationary noise, age-dependent reference data are available, and our results closely match those previously reported by O’Brien et al. (2024). Both studies show comparable median/mean values across age groups between 40 and 69 years, with O’Brien et al. (2024) reporting a mean SRT of -6.3 ± 1.1 dB SNR reported for the age group 40-44 years and -6.1 ± 1.1 dB SNR for the age group 45-49 years, while this study estimated a median SRT of -6.3 dB SNR (IQR [-6.9, -5.8] dB SNR; cf. Table A2) for the age group 40-49 years. The slope is highly comparable as well, with sex-specific values of 0.64 dB per age decade for men and 0.54 dB per age decade for women (O’Brien et al., 2024), and 0.60 dB per age decade in our study (cf. Table 1).

For the TIN test, both frequencies showed a significant age effect, although no directly comparable literature data exist. The observed age-related patterns were similar to those found for the Matrix test, although less pronounced in terms of correlation strength and significance level. This similarity is plausible, as TIN is expected to capture similar suprathreshold auditory processing deficits (Schädler et al., 2020; Hülsmeier and Kollmeier, 2022). Our data generally support this hypothesis, but the variability across participants was substantially higher for TIN than for the Matrix test. A similar pattern of pronounced inter-individual variability in TIN thresholds at higher frequencies has also been reported in older listeners with clinically normal hearing (Ralli et al., 2018), underscoring that high-frequency TIN detection is particularly susceptible to age-related processing differences. Future studies should investigate whether this variability primarily reflects genuine individual listener differences or whether it is partly driven by differences in test–retest reliability across the applied measures.

For ECochG, the significant age effect for wave I latency was expected (Konrad-Martin et al., 2012; Mehraei et al., 2016; Johannesen et al., 2019). Relating the increasing latency of wave I to the stable interpeak latencies I-V and III-V indicates a global shift of the auditory brainstem response to longer latencies. This pattern suggests delayed neural transmission at the auditory nerve level, with preserved relative transmission along the auditory pathway. Importantly, wave I latency did not correlate significantly with PTA, indicating that the effect is not merely driven by reduced audibility or cochlear dysfunction. Instead, the absence of a PTA dependency supports the interpretation of a predominantly neural origin of the latency increase. Several mechanisms may account for this pattern. Previous studies have reported similar findings, showing prolonged wave I latencies and reduced amplitudes in older adults with clinically normal hearing thresholds (Konrad-Martin et al., 2012; Mehraei et al., 2016; Johannesen et al., 2019). These results have been linked to age-related cochlear synaptopathy, auditory-nerve fiber loss, and demyelination, which collectively reduce neural synchrony and conduction velocity. Such mechanisms can delay the generation of wave I while leaving interpeak intervals largely unaffected—consistent with the pattern observed in our data.

For vestibular tests, age-related dependencies on amplitudes would have been expected, such as decreasing SPV in the caloric test (particularly for warm irrigation), and a reduction in VEMP interpeak amplitude in a normal-hearing population (Maes et al., 2010). In contrast, the data of the present paper showed only a weak trend for decreasing oVEMP normalised amplitude, which is not directly comparable to the interpeak amplitude due to different EMG potentials across participants. For vHIT gains, the absence of age-dependent changes was consistent with previous findings of McGarvie et al. (2015). For all vestibular tests, no age dependencies were expected for the asymmetry measures (Maes et al., 2010; McGarvie et al., 2015), which is plausible as both sides are likely to be similarly affected by the aging process.

### Establishing reference data for the proposed test battery

Taken together, the presented data establish a comprehensive, cross-domain source of age-dependent normative data for the proposed test battery (objective 4). As discussed above, most identified effects correspond well to findings from the literature, but even more importantly, for many tests age-dependent normative data have so far been lacking, and corresponding expectations remain unclear. To address this gap, we provide age-dependent reference data for the better-hearing half of the population, defined as participants with PTA better than the age-dependent median according to ISO 7029 (2017). The outcomes of this study are presented, both graphically (Figures 2-11) and statistically (Table 1), and summarised as median values and interquartile ranges per age decade in the supplementary Table A2. These reference data are important for interpreting data collected on hearing-impaired participants. Identified dependencies on age or PTA should be explicitly considered to ensure accurate interpretation and be used as covariates in future statistical analyses. Conversely, tests for which no age or PTA effects were identified can be directly interpreted with respect to the auditory or vestibular functions they characterise, as they likely provide complementary, suprathreshold information. Such tests are particularly valuable for revealing subclinical or central processing alterations beyond audibility. Examples for this can be found in the included domains of audiological-functional, audiological-physiological, and vestibular-physiological tests. As a parameter derived from ACALOS, the binaural loudness summation (BLS) was found to be age- and PTA-independent. The observed value range between 0 and 20 dB in our study is consistent with previous studies (Oetting et al., 2016; van Beurden et al., 2018), which reported median BLS values for normal-hearing participants of about 10 dB, and a comparably large interindividual variability. This variability highlights the diversity of individual auditory characteristics and hence underscores the diagnostic potential of BLS as a measure of binaural processing.

In contrast to the other tests, otoacoustic emissions were only investigated in terms of presence or absence. Statistical analysis of the binary response pattern with respect to age and PTA was not conducted due to the very low number of participants with absent responses (8 for TEOAE and 5 for DPOAE), which would limit the interpretation. Absent responses were generally found for participants older than 60 years. This finding is in line with Stover & Norton (1993), who found increasing absence of otoacoustic emissions with increasing age in normal-hearing participants. However, they state that this effect is likely not independent from PTA; both variables have to be considered for interpretation. DPOAEs can serve as a valuable instrument to either confirm or exclude the presence of hearing loss, and can be useful for interpreting electrocochleography (ECochG) results.

### Limitations

The results of this study should be interpreted in the context of certain limitations. First, the relatively small number of French participants influenced the centre effect analysis. To provide a fair and statistically sound comparison between French and German data, additional analysis was required to match the two groups based on age and PTA. Despite the limited sample size, the overall consistency of findings across centres supports the validity of the observed effects and interpretations. Nevertheless, a larger French group would likely further reduce the residual centre effects, thereby enhancing confidence in the multicentre comparability of the test battery. Second, three participants (two at UOL and one at IDA) were included in the study despite reporting diabetes type 2 in the inclusion questionnaire, but were retained in the analysis, as their data did not differ notably from those of other participants. Future studies might consider stricter exclusion criteria or targeted subgroup analyses to account for potential metabolic influences on auditory or vestibular measures. Third, data collection was performed in the scope of a larger project, which will include hearing-impaired listeners. Consequently, repeated measurements were not obtained in the present age-adjusted normal-hearing cohort, and test-retest reliability for some tests remains unclear. Future work should therefore include dedicated repeatability assessments to better quantify measurement precision and to relate test–retest reliability to the observed interindividual variability, as discussed above.

A more general methodological limitation concerns variability in data export functionalities and documentation across the commercial measurement devices employed. This relates to differences in available data formats, access to raw data, and the degree of transparency regarding internal signal processing (for example, information about which trials were considered as outliers and consequently removed from the calculation of averaged output parameters). Full reproducibility requires access to raw data, particularly for cross-device or cross-centre comparisons. For instance, in this study, asymmetry indices for vHIT and VEMP were recalculated from the raw signals to mitigate potential inconsistencies in device-specific averaging algorithms, illustrating a practical approach to improving data comparability.

### Future work

The present study focused on analysing separate PTA and age dependencies in participants with age-normal PTA, as a first step towards characterising the proposed cross-domain test battery and establishing age-dependent normative data. Building on this foundation, subsequent work will explore relationships between the different tests within the test battery, including across the domains of audiological-functional, audiological-physiological, and vestibular-physiological tests. Examining these interrelations, even within the present control group, will provide valuable insight into the extent to which the various tests capture complementary or overlapping auditory and vestibular processes.

In the next phase, data from hearing-impaired participants will be compared against the reference data presented here, enabling investigation of a broad range of research questions in the context of the PRESAGE project and beyond. For example, these analyses will help clarify the contribution of individual tests to the characterisation and identification of (untimely) presbycusis, and may support the development of an optimised, clinically efficient test battery. Combining data across all age groups and the complete PTA range will further allow the exploration of multivariate patterns within the dataset. This could be achieved, for example, through unsupervised clustering methods (e.g., Saak et al., 2022; 2025; Sanchez-Lopez et al., 2018). Given that several participants from the age-adjusted normal-hearing cohort already exhibited abnormal results in some tests, these analyses may reveal which test combinations most effectively differentiate between subclinical or emerging forms of dysfunction beyond audibility. To support open science and reproducibility, the data presented in this study will later be published in an open repository together with data from measurements with hearing-impaired participants.

An additional future direction involves integrating genetic information with the audiological and vestibular phenotypes. As part of the ongoing PRESAGE study, blood samples are being collected to enable genotype–phenotype correlation analyses. This approach will allow examination of whether specific audiological-vestibular test patterns correspond to known or novel (monogenic) variants associated with hearing loss. Such work would extend the findings of Boucher et al. (2020), who identified genetic forms of hearing loss based solely on audiometric data, by incorporating a much richer set of phenotypic measures.

Finally, the harmonisation and standardisation of audiological and vestibular data are essential for advancing multicentre research. Establishing shared data standards will support unified data formats and complete documentation of test procedures and measurement conditions (Vercammen et al., 2025). These efforts will facilitate reproducibility, interoperability across devices and centres, and open data sharing in future large-scale studies.

## Conclusions

The main findings of this study are:

- Comparable results across sites can be obtained with the proposed test battery; detailed information about employed equipment and test conditions has to be available to assess and achieve comparability, emphasising the need for complete information and metadata.
- Within the group of participants with age-normal PTA, a few PTA effects were identified for the Matrix test and otoacoustic emissions. Additionally, several tests, particularly in the vestibular domain, showed high variability not directly related to PTA. These findings emphasise the need to account for audibility-related dependencies when interpreting cross-domain measures, even among participants with clinically normal hearing.
- Age effects were found for all audiological-functional tests and single variables of audiological-physiological tests, but not for vestibular tests.
- Our data provide important reference data to be taken into account as covariates (PTA, age, or both) for interpretation of the respective tests, and to compare data for hearing-impaired participants to these reference data.
- Future work provides the opportunity to establish correlations and further relationships between the tests of the audiological-vestibular test battery, on the group considered here as well as on data for hearing-impaired participants with PTA higher than the age-dependent median, or even the highest 10% per age (untimely age-related hearing loss).

## Data Availability

Data are planned to be published in an open repository together with the hearing-impaired data at a later stage of the project. Data are available upon motivated request. The full reproducible analysis code is available on Zenodo.

https://doi.org/10.5281/zenodo.17950319

## Acknowledgements

The authors thank Thinhinane Arris, Simona Caldani, Marta Campi, Clément Gaultier, Doris Medina Garin, and Anita Gorges for their contributions to data collection. The authors also thank Céline Quinsac, Madalina Alves Ferreira, Annette Schult, Keno Sprenger, and Anita Gorges for management and administrative support. The authors thank the Clinical Research Coordination Office (CRCO) of the Institut Pasteur for their help with biomedical regulatory and ethical aspects of the project.

## Declaration of conflicting interests

AC is employed by Sonova Audiological Care France. The authors declare that this affiliation did not influence study design, data collection and analysis, decision to publish, or preparation of the manuscript.

## Data availability statement

Data are planned to be published in an open repository together with the hearing-impaired data at a later stage of the project. Data are available upon motivated request. The full reproducible analysis code is available on Zenodo (https://doi.org/10.5281/zenodo.17950319).

## Funding

This work was funded by a French-German grant by the Agence Nationale de la Recherche (ANR) and the German Research Foundation (DFG), project numbers ANR-21-CE14-0075 and 490819095. This work was supported by a grant from Fondation Pour l’Audition (FPA) to the CERIAH (FPA IDA10). This work has benefited from a French government grant managed by the Agence Nationale de la Recherche under the France 2030 program, reference ANR-23-IAHU-0003.

## Author contributions

**MB**: Conceptualisation, Methodology, Software, Validation, Formal Analysis, Visualisation, Data Collection, Data Curation, Writing – Original Draft, Writing – Review & Editing; **SK**: Conceptualisation, Methodology, Software, Validation, Formal Analysis, Visualisation, Data Collection, Data Curation, Writing – Original Draft, Writing – Review & Editing; **ASM**: Data Collection, Data Curation, Writing – Original Draft, Writing – Review & Editing; **KK**: Data Collection, Data Curation, Writing – Original Draft, Writing – Review & Editing; **SH**: Data Collection, Data Curation, Writing – Review & Editing; **EP**: Validation, Data Collection, Data Curation, Writing – Review & Editing; **AC**: Data Collection, Writing – Review & Editing; **AR**: Writing – Review & Editing, Funding acquisition; **HTV**: Writing – Review & Editing, Funding acquisition; **SVW**: Data Collection, Writing – Review & Editing; **GG**: Validation, Formal Analysis, Data Collection, Data Curation, Validation, Writing – Review & Editing; **AW**: Conceptualisation, Validation, Writing – Original Draft, Writing – Review & Editing; **PA**: Conceptualisation, Writing – Review & Editing, Funding acquisition; **BK**: Conceptualisation, Writing – Review & Editing, Funding acquisition.

All authors have read and agreed to the final version of the manuscript.

## APPENDIX

**Table A1.** Centre effect analysis summary for the matched French (IDA) and German (UOL) subset. Shown are results from Levene’s tests assessing the assumption of homogeneity of variance, and generalised linear models (GLMs), adjusted for age and PTA. Centre was coded with IDA as the reference level. Significant p-values (p<0.05) are shown in bold.

| Test / Variable | Levene's test |  |  |  | Centre effect (GLM) |  |  |  |  |
| --- | --- | --- | --- | --- | --- | --- | --- | --- | --- |
| | n<sub>UOL</sub> | n<sub>IDA</sub> | F | p | $\beta_1$ | SE | adj. R<sup>2</sup> | p | Cohen's d |
| **ACALOS** |  |  |  |  |  |  |  |  |  |
| L15 [dB SPL] | 16 | 16 | 0.997 | 0.3261 | -5.138 | 1.811 | 0.281 | **0.008** | -0.937 |
| L35 [dB SPL] | 16 | 16 | 7.131 | **0.0121** | -5.078 | 2.027 | 0.151 | **0.018** | -0.938 |
| DR [dB] | 16 | 16 | 0.005 | 0.9434 | 0.060 | 1.543 | 0.024 | 0.969 | -0.035 |
| BLS [dB] | 16 | 16 | 4.648 | 0.0392 | 1.143 | 1.807 | 0.04 | 0.532 | 0.126 |
| **TIN** |  |  |  |  |  |  |  |  |  |
| ERB<sub>500Hz</sub> [dB SNR] | 16 | 13 | 0.058 | 0.8113 | 0.771 | 0.668 | -0.022 | 0.259 | 0.436 |
| ERB<sub>2000Hz</sub> [dB SNR] | 16 | 13 | 0.383 | 0.5413 | 0.170 | 1.000 | -0.081 | 0.867 | 0.04 |
| **Matrix test** |  |  |  |  |  |  |  |  |  |
| SRT SSN [dB SNR] | 16 | 16 | 10.526 | **0.0029** | 1.493 | 0.456 | 0.425 | **0.003** | 1.057 |
| SRT ICRA5-250 [dB SNR] | 16 | 16 | 0.034 | 0.8546 | 3.178 | 0.799 | 0.673 | **0.000** | 1.053 |
| SRT<sub>norm</sub> SSN [dB SNR] | 16 | 16 | 10.526 | 0.0029 | 0.393 | 0.456 | 0.275 | 0.396 | 0.325 |
| SRT<sub>norm</sub> ICRA5-250 [dB SNR] | 16 | 16 | 0.034 | 0.8546 | -0.022 | 0.799 | 0.579 | 0.979 | 0.136 |
| **MEMR** |  |  |  |  |  |  |  |  |  |
| Average threshold [dB SPL] | 12 | 15 | 0.012 | 0.9144 | 0.749 | 2.088 | -0.065 | 0.723 | 0.19 |
| **ECochG** |  |  |  |  |  |  |  |  |  |
| Latency wave I [ms]<sup>1</sup> | 16 | 10 | 0.118 | 0.7341 | 0.040 | 0.048 | 0.167 | 0.423 | 0.175 |
| Interpeak I-V [ms]<sup>1</sup> | 16 | 10 | 0.61 | 0.4424 | -0.153 | 0.077 | 0.111 | 0.061 | -0.877 |
| Interpeak III-V [ms]<sup>1</sup> | 16 | 14 | 1.757 | 0.1957 | -0.093 | 0.069 | -0.014 | 0.192 | -0.477 |
| **Caloric test** |  |  |  |  |  |  |  |  |  |
| SPV cold [°/s]<sup>2</sup> | 14 | 15 | 3.334 | 0.0789 | 7.785 | 2.966 | 0.231 | **0.015** | 0.861 |
| SPV warm [°/s] | 14 | 14 | 0.323 | 0.5747 | 6.638 | 5.785 | 0.016 | 0.263 | 0.37 |
| UW [%]<sup>2</sup> | 14 | 14 | 3.587 | 0.0694 | 10.167 | 6.448 | 0.12 | 0.128 | 0.72 |
| DP [%] | 14 | 14 | 1.158 | 0.2917 | 3.424 | 5.824 | 0.108 | 0.562 | 0.118 |
| **vHIT** |  |  |  |  |  |  |  |  |  |
| Gain Anterior | 15 | 16 | 5.238 | **0.0296** | 0.033 | 0.045 | -0.062 | 0.476 | 0.228 |
| Gain Lateral | 15 | 16 | 0.703 | 0.4086 | -0.124 | 0.041 | 0.277 | **0.006** | -1.088 |
| Gain Posterior | 15 | 16 | 0.583 | 0.4513 | 0.037 | 0.040 | 0.121 | 0.370 | 0.202 |
| AR Anterior [%] | 15 | 16 | 10.476 | **0.003** | -3.702 | 3.003 | -0.021 | 0.228 | -0.409 |
| AR Lateral [%] | 15 | 16 | 3.428 | 0.0743 | -1.228 | 1.592 | -0.062 | 0.447 | -0.301 |
| AR Posterior [%] | 14 | 16 | 1.538 | 0.2252 | 0.770 | 2.182 | -0.024 | 0.727 | 0.108 |
| **cVEMP** |  |  |  |  |  |  |  |  |  |
| EMG-normalised P-N amplitude [μV] | 14 | 15 | 3.566 | 0.0698 | 0.354 | 0.162 | 0.075 | **0.038** | 0.818 |
| AR [%] | 13 | 15 | 4.548 | **0.0426** | 4.861 | 7.696 | -0.093 | 0.534 | 0.252 |
| **oVEMP** |  |  |  |  |  |  |  |  |  |
| EMG-normalised P-N amplitude [μV] | 14 | 11 | 0.773 | 0.3883 | 0.560 | 0.140 | 0.356 | **0.001** | 1.699 |
| AR [%] | 14 | 9 | 2.381 | 0.1378 | 1.566 | 10.460 | 0.007 | 0.883 | 0.186 |

<sup>1</sup>only latencies measured at 115 dB SPL were included; <sup>2</sup>monothermal SPVs were excluded.

**Table A2:**
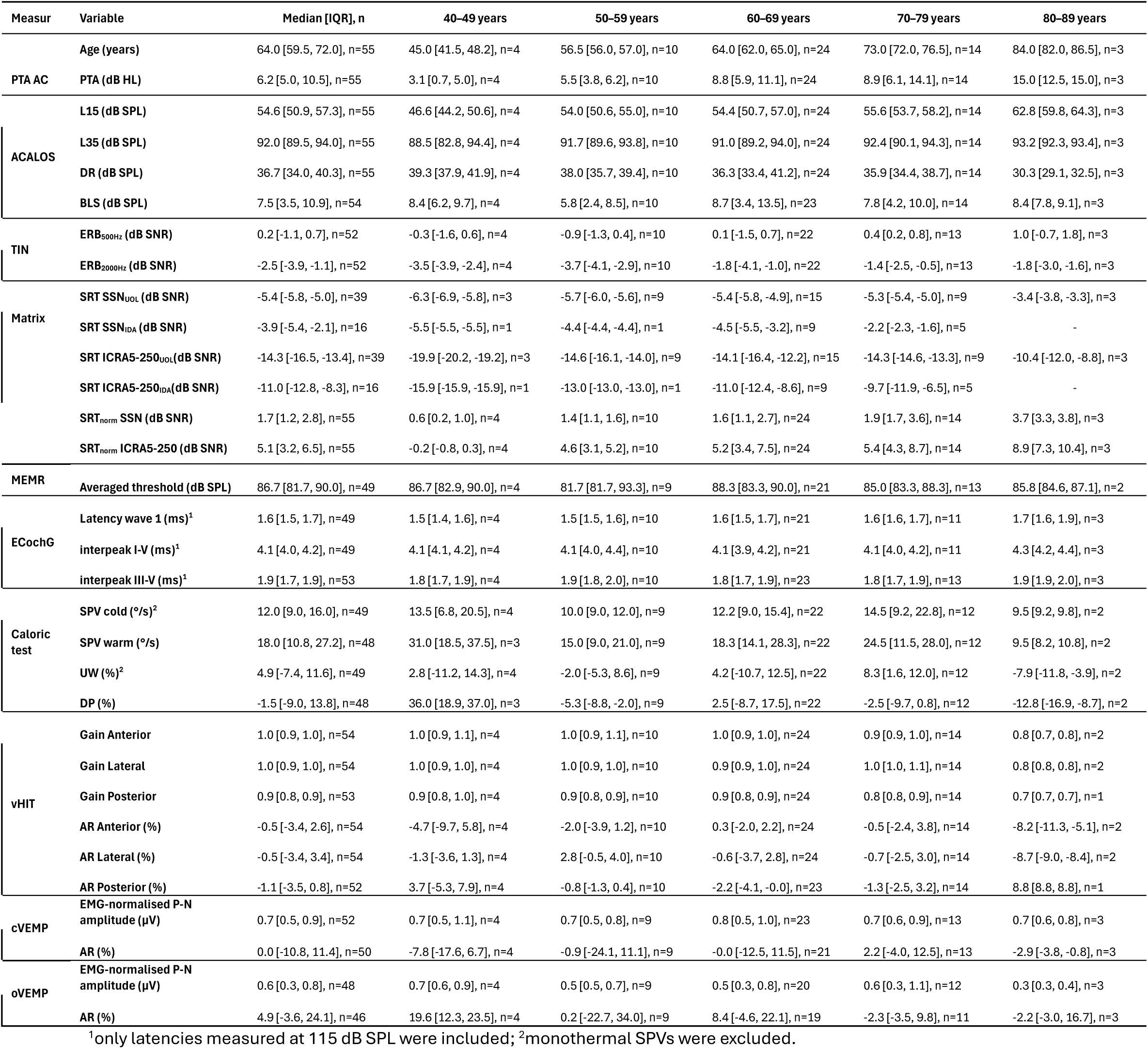
PRESAGE test battery results for normal-hearing participants, shown as median, [IQR] with sample size (n) for the combined dataset and by age group.

| Measur | Variable | Median [IQR], n | 40–49 years | 50–59 years | 60–69 years | 70–79 years | 80–89 years |
| --- | --- | --- | --- | --- | --- | --- | --- |
| PTA AC | Age (years) | 64.0 [59.5, 72.0], n=55 | 45.0 [41.5, 48.2], n=4 | 56.5 [56.0, 57.0], n=10 | 64.0 [62.0, 65.0], n=24 | 73.0 [72.0, 76.5], n=14 | 84.0 [82.0, 86.5], n=3 |
|  | PTA (dB HL) | 6.2 [5.0, 10.5], n=55 | 3.1 [0.7, 5.0], n=4 | 5.5 [3.8, 6.2], n=10 | 8.8 [5.9, 11.1], n=24 | 8.9 [6.1, 14.1], n=14 | 15.0 [12.5, 15.0], n=3 |
| ACALOS | L15 (dB SPL) | 54.6 [50.9, 57.3], n=55 | 46.6 [44.2, 50.6], n=4 | 54.0 [50.6, 55.0], n=10 | 54.4 [50.7, 57.0], n=24 | 55.6 [53.7, 58.2], n=14 | 62.8 [59.8, 64.3], n=3 |
|  | L35 (dB SPL) | 92.0 [89.5, 94.0], n=55 | 88.5 [82.8, 94.4], n=4 | 91.7 [89.6, 93.8], n=10 | 91.0 [89.2, 94.0], n=24 | 92.4 [90.1, 94.3], n=14 | 93.2 [92.3, 93.4], n=3 |
|  | DR (dB SPL) | 36.7 [34.0, 40.3], n=55 | 39.3 [37.9, 41.9], n=4 | 38.0 [35.7, 39.4], n=10 | 36.3 [33.4, 41.2], n=24 | 35.9 [34.4, 38.7], n=14 | 30.3 [29.1, 32.5], n=3 |
|  | BLS (dB SPL) | 7.5 [3.5, 10.9], n=54 | 8.4 [6.2, 9.7], n=4 | 5.8 [2.4, 8.5], n=10 | 8.7 [3.4, 13.5], n=23 | 7.8 [4.2, 10.0], n=14 | 8.4 [7.8, 9.1], n=3 |
| TIN | ERB<sub>500Hz</sub> (dB SNR) | 0.2 [-1.1, 0.7], n=52 | -0.3 [-1.6, 0.6], n=4 | -0.9 [-1.3, 0.4], n=10 | 0.1 [-1.5, 0.7], n=22 | 0.4 [0.2, 0.8], n=13 | 1.0 [-0.7, 1.8], n=3 |
|  | ERB<sub>2000Hz</sub> (dB SNR) | -2.5 [-3.9, -1.1], n=52 | -3.5 [-3.9, -2.4], n=4 | -3.7 [-4.1, -2.9], n=10 | -1.8 [-4.1, -1.0], n=22 | -1.4 [-2.5, -0.5], n=13 | -1.8 [-3.0, -1.6], n=3 |
| Matrix | SRT SSN<sub>UOL</sub> (dB SNR) | -5.4 [-5.8, -5.0], n=39 | -6.3 [-6.9, -5.8], n=3 | -5.7 [-6.0, -5.6], n=9 | -5.4 [-5.8, -4.9], n=15 | -5.3 [-5.4, -5.0], n=9 | -3.4 [-3.8, -3.3], n=3 |
|  | SRT SSN<sub>IDA</sub> (dB SNR) | -3.9 [-5.4, -2.1], n=16 | -5.5 [-5.5, -5.5], n=1 | -4.4 [-4.4, -4.4], n=1 | -4.5 [-5.5, -3.2], n=9 | -2.2 [-2.3, -1.6], n=5 | - |
|  | SRT ICRA5-250<sub>UOL</sub> (dB SNR) | -14.3 [-16.5, -13.4], n=39 | -19.9 [-20.2, -19.2], n=3 | -14.6 [-16.1, -14.0], n=9 | -14.1 [-16.4, -12.2], n=15 | -14.3 [-14.6, -13.3], n=9 | -10.4 [-12.0, -8.8], n=3 |
|  | SRT ICRA5-250<sub>IDA</sub> (dB SNR) | -11.0 [-12.8, -8.3], n=16 | -15.9 [-15.9, -15.9], n=1 | -13.0 [-13.0, -13.0], n=1 | -11.0 [-12.4, -8.6], n=9 | -9.7 [-11.9, -6.5], n=5 | - |
|  | SRT<sub>norm</sub> SSN (dB SNR) | 1.7 [1.2, 2.8], n=55 | 0.6 [0.2, 1.0], n=4 | 1.4 [1.1, 1.6], n=10 | 1.6 [1.1, 2.7], n=24 | 1.9 [1.7, 3.6], n=14 | 3.7 [3.3, 3.8], n=3 |
|  | SRT<sub>norm</sub> ICRA5-250 (dB SNR) | 5.1 [3.2, 6.5], n=55 | -0.2 [-0.8, 0.3], n=4 | 4.6 [3.1, 5.2], n=10 | 5.2 [3.4, 7.5], n=24 | 5.4 [4.3, 8.7], n=14 | 8.9 [7.3, 10.4], n=3 |
| MEMR | Averaged threshold (dB SPL) | 86.7 [81.7, 90.0], n=49 | 86.7 [82.9, 90.0], n=4 | 81.7 [81.7, 93.3], n=9 | 88.3 [83.3, 90.0], n=21 | 85.0 [83.3, 88.3], n=13 | 85.8 [84.6, 87.1], n=2 |
| ECochG | Latency wave 1 (ms)<sup>1</sup> | 1.6 [1.5, 1.7], n=49 | 1.5 [1.4, 1.6], n=4 | 1.5 [1.5, 1.6], n=10 | 1.6 [1.5, 1.7], n=21 | 1.6 [1.6, 1.7], n=11 | 1.7 [1.6, 1.9], n=3 |
|  | interpeak I-V (ms)<sup>1</sup> | 4.1 [4.0, 4.2], n=49 | 4.1 [4.1, 4.2], n=4 | 4.1 [4.0, 4.4], n=10 | 4.1 [3.9, 4.2], n=21 | 4.1 [4.0, 4.2], n=11 | 4.3 [4.2, 4.4], n=3 |
|  | interpeak III-V (ms)<sup>1</sup> | 1.9 [1.7, 1.9], n=53 | 1.8 [1.7, 1.9], n=4 | 1.9 [1.8, 2.0], n=10 | 1.8 [1.7, 1.9], n=23 | 1.8 [1.7, 1.9], n=13 | 1.9 [1.9, 2.0], n=3 |
| Caloric test | SPV cold (°/s)<sup>2</sup> | 12.0 [9.0, 16.0], n=49 | 13.5 [6.8, 20.5], n=4 | 10.0 [9.0, 12.0], n=9 | 12.2 [9.0, 15.4], n=22 | 14.5 [9.2, 22.8], n=12 | 9.5 [9.2, 9.8], n=2 |
|  | SPV warm (°/s) | 18.0 [10.8, 27.2], n=48 | 31.0 [18.5, 37.5], n=3 | 15.0 [9.0, 21.0], n=9 | 18.3 [14.1, 28.3], n=22 | 24.5 [11.5, 28.0], n=12 | 9.5 [8.2, 10.8], n=2 |
|  | UW (%)<sup>2</sup> | 4.9 [-7.4, 11.6], n=49 | 2.8 [-11.2, 14.3], n=4 | -2.0 [-5.3, 8.6], n=9 | 4.2 [-10.7, 12.5], n=22 | 8.3 [1.6, 12.0], n=12 | -7.9 [-11.8, -3.9], n=2 |
|  | DP (%) | -1.5 [-9.0, 13.8], n=48 | 36.0 [18.9, 37.0], n=3 | -5.3 [-8.8, -2.0], n=9 | 2.5 [-8.7, 17.5], n=22 | -2.5 [-9.7, 0.8], n=12 | -12.8 [-16.9, -8.7], n=2 |
| vHIT | Gain Anterior | 1.0 [0.9, 1.0], n=54 | 1.0 [0.9, 1.1], n=4 | 1.0 [0.9, 1.1], n=10 | 1.0 [0.9, 1.0], n=24 | 0.9 [0.9, 1.0], n=14 | 0.8 [0.7, 0.8], n=2 |
|  | Gain Lateral | 1.0 [0.9, 1.0], n=54 | 1.0 [0.9, 1.0], n=4 | 1.0 [0.9, 1.0], n=10 | 0.9 [0.9, 1.0], n=24 | 1.0 [1.0, 1.1], n=14 | 0.8 [0.8, 0.8], n=2 |
|  | Gain Posterior | 0.9 [0.8, 0.9], n=53 | 0.9 [0.8, 1.0], n=4 | 0.9 [0.8, 0.9], n=10 | 0.9 [0.8, 0.9], n=24 | 0.8 [0.8, 0.9], n=14 | 0.7 [0.7, 0.7], n=1 |
|  | AR Anterior (%) | -0.5 [-3.4, 2.6], n=54 | -4.7 [-9.7, 5.8], n=4 | -2.0 [-3.9, 1.2], n=10 | 0.3 [-2.0, 2.2], n=24 | -0.5 [-2.4, 3.8], n=14 | -8.2 [-11.3, -5.1], n=2 |
|  | AR Lateral (%) | -0.5 [-3.4, 3.4], n=54 | -1.3 [-3.6, 1.3], n=4 | 2.8 [-0.5, 4.0], n=10 | -0.6 [-3.7, 2.8], n=24 | -0.7 [-2.5, 3.0], n=14 | -8.7 [-9.0, -8.4], n=2 |
|  | AR Posterior (%) | -1.1 [-3.5, 0.8], n=52 | 3.7 [-5.3, 7.9], n=4 | -0.8 [-1.3, 0.4], n=10 | -2.2 [-4.1, -0.0], n=23 | -1.3 [-2.5, 3.2], n=14 | 8.8 [8.8, 8.8], n=1 |
| cVEMP | EMG-normalised P-N amplitude (μV) | 0.7 [0.5, 0.9], n=52 | 0.7 [0.5, 1.1], n=4 | 0.7 [0.5, 0.8], n=9 | 0.8 [0.5, 1.0], n=23 | 0.7 [0.6, 0.9], n=13 | 0.7 [0.6, 0.8], n=3 |
|  | AR (%) | 0.0 [-10.8, 11.4], n=50 | -7.8 [-17.6, 6.7], n=4 | -0.9 [-24.1, 11.1], n=9 | -0.0 [-12.5, 11.5], n=21 | 2.2 [-4.0, 12.5], n=13 | -2.9 [-3.8, -0.8], n=3 |
| oVEMP | EMG-normalised P-N amplitude (μV) | 0.6 [0.3, 0.8], n=48 | 0.7 [0.6, 0.9], n=4 | 0.5 [0.5, 0.7], n=9 | 0.5 [0.3, 0.8], n=20 | 0.6 [0.3, 1.1], n=12 | 0.3 [0.3, 0.4], n=3 |
|  | AR (%) | 4.9 [-3.6, 24.1], n=46 | 19.6 [12.3, 23.5], n=4 | 0.2 [-22.7, 34.0], n=9 | 8.4 [-4.6, 22.1], n=19 | -2.3 [-3.5, 9.8], n=11 | -2.2 [-3.0, 16.7], n=3 |

<sup>1</sup>only latencies measured at 115 dB SPL were included; <sup>2</sup>monothermal SPVs were excluded.

## Footnotes

^1^http://medi.uni-oldenburg.de/afc

## Notes

### Author Declarations

The pre-medical Ethics Committee of the Carl von Ossietzky Universitaet Oldenburg (2022-074) and the Comite de Protection des Personnes (CPP; Committee for the protection of persons) Sud Ouest et Outre-mer 1 (2021-062) gave ethical approval for this work.

